# SARS-CoV-2 incidence, transmission and reinfection in a rural and an urban setting: results of the PHIRST-C cohort study, South Africa, 2020-2021

**DOI:** 10.1101/2021.07.20.21260855

**Authors:** Cheryl Cohen, Jackie Kleynhans, Anne von Gottberg, Meredith L McMorrow, Nicole Wolter, Jinal N. Bhiman, Jocelyn Moyes, Mignon du Plessis, Maimuna Carrim, Amelia Buys, Neil A Martinson, Kathleen Kahn, Stephen Tollman, Limakatso Lebina, Floidy Wafawanaka, Jacques du Toit, Francesc Xavier Gómez-Olivé, Fatimah S. Dawood, Thulisa Mkhencele, Kaiyun Sun, Cécile Viboud, for the PHIRST group, Stefano Tempia

**Affiliations:** Centre for Respiratory Diseases and Meningitis, National Institute for Communicable Diseases of the National Health Laboratory Service, Johannesburg, South Africa; School of Public Health, Faculty of Health Sciences, University of the Witwatersrand, Johannesburg, South Africa; Influenza Division, Centers for Disease Control and Prevention, Atlanta, Georgia, United States of America; School of Pathology, Faculty of Health Sciences, University of the Witwatersrand, Johannesburg, South Africa; Perinatal HIV Research Unit, MRC Soweto Matlosana Collaborating Centre for HIV/AIDS and TB, University of the Witwatersrand, South Africa; DST/NRF Centre of Excellence for Biomedical Tuberculosis Research, University of the Witwatersrand, Johannesburg, South Africa; Johns Hopkins University Center for TB Research, Baltimore, Maryland, United States of America; MRC/Wits Rural Public Health and Health Transitions Research Unit (Agincourt), Faculty of Health Sciences, School of Public Health, University of the Witwatersrand, Johannesburg, South Africa; Africa Health Research Institute, KwaZulu-Natal, South Africa; Division of International Epidemiology and Population Studies, Fogarty International Center, National Institutes of Health, Bethesda, MD, USA

**Keywords:** SARS-CoV-2, burden, transmission, household, South Africa, HIV, rural, urban

## Abstract

**Background:** By August 2021, South Africa experienced three SARS-CoV-2 waves; the second and third associated with emergence of Beta and Delta variants respectively.

**Methods:** We conducted a prospective cohort study during July 2020-August 2021 in one rural and one urban community. Mid-turbinate nasal swabs were collected twice-weekly from household members irrespective of symptoms and tested for SARS-CoV-2 using real-time reverse transcription polymerase chain reaction (rRT-PCR). Serum was collected every two months and tested for anti-SARS-CoV-2 antibodies.

**Results:** Among 115,759 nasal specimens from 1,200 members (follow-up rate 93%), 1976 (2%) were SARS-CoV-2-positive. By rRT-PCR and serology combined, 62% (749/1200) of individuals experienced ≥1 SARS-CoV-2 infection episode, and 12% (87/749) experienced reinfection. Of 662 PCR-confirmed episodes with available data, 15% (n=97) were associated with ≥1 symptom. Among 222 households, 200 (90%) had ≥1 SARS-CoV-2-positive individual. Household cumulative infection risk (HCIR) was 25% (213/856). On multivariable analysis, accounting for age and sex, index case lower cycle threshold value (OR 3.9, 95%CI 1.7-8.8), urban community (OR 2.0,95%CI 1.1-3.9), Beta (OR 4.2, 95%CI 1.7-10.1) and Delta (OR 14.6, 95%CI 5.7-37.5) variant infection were associated with increased HCIR. HCIR was similar for symptomatic (21/110, 19%) and asymptomatic (195/775, 25%) index cases (p=0.165). Attack rates were highest in individuals aged 13-18 years and individuals in this age group were more likely to experience repeat infections and to acquire SARS-CoV-2 infection. People living with HIV who were not virally supressed were more likely to develop symptomatic illness, and shed SARS-CoV-2 for longer compared to HIV-uninfected individuals.

**Conclusions:** In this study, 85% of SARS-CoV-2 infections were asymptomatic and index case symptom status did not affect HCIR, suggesting a limited role for control measures targeting symptomatic individuals. Increased household transmission of Beta and Delta variants, likely contributed to successive waves, with >60% of individuals infected by the end of follow-up.

**Research in context:** *Evidence before this study:* Previous studies have generated wide-ranging estimates of the proportion of SARS-CoV-2 infections which are asymptomatic. A recent systematic review found that 20% (95% CI 3%-67%) of severe acute respiratory syndrome coronavirus 2 (SARS-CoV-2) infections remained asymptomatic throughout infection and that transmission from asymptomatic individuals was reduced. A systematic review and meta-analysis of 87 household transmission studies of SARS-CoV-2 found an estimated secondary attack rate of 19% (95% CI 16-22). The review also found that household secondary attack rates were increased from symptomatic index cases and that adults were more likely to acquire infection. As of December 2021, South Africa experienced three waves of SARS-CoV-2 infections; the second and third waves were associated with circulation of Beta and Delta variants respectively. SARS-CoV-2 vaccines became available in February 2021, but uptake was low in study sites reaching 5% fully vaccinated at the end of follow up. Studies to quantify the burden of asymptomatic infections, symptomatic fraction, reinfection frequency, duration of shedding and household transmission of SARS-CoV-2 from asymptomatically infected individuals have mostly been conducted as part of outbreak investigations or in specific settings. Comprehensive systematic community studies of SARS-CoV-2 burden and transmission including for the Beta and Delta variants are lacking, especially in low vaccination settings.

*Added value of this study:* We conducted a unique detailed COVID-19 household cohort study over a 13 month period in South Africa, with real time reverse transcriptase polymerase chain reaction (rRT-PCR) testing twice a week irrespective of symptoms and bimonthly serology. By the end of the study in August 2021, 749 (62%) of 1200 individuals from 222 randomly sampled households in a rural and an urban community in South Africa had at least one confirmed SARS-CoV-2 infection, detected on rRT-PCR and/or serology, and 12% (87/749) experienced reinfection. Symptom data were analysed for 662 rRT-PCR-confirmed infection episodes that occurred >14 days after the start of follow-up (of a total of 718 rRT-PCR-confirmed episodes), of these, 15% (n=97) were associated with one or more symptoms. Among symptomatic indvidiausl, 9% (n=9) were hospitalised and 2% (n=2) died. Ninety percent (200/222) of included households, had one or more individual infected with SARS-CoV-2 on rRT-PCR and/or serology within the household. SARS-CoV-2 infected index cases transmitted the infection to 25% (213/856) of susceptible household contacts. Index case ribonucleic acid (RNA) viral load proxied by rRT-PCR cycle threshold value was strongly predictive of household transmission. Presence of symptoms in the index case was not associated with household transmission. Household transmission was four times greater from index cases infected with Beta variant and fifteen times greater from index cases infected with Delta variant compared to wild-type infection. Attack rates were highest in individuals aged 13-18 years and individuals in this age group were more likely to experience repeat infections and to acquire SARS-CoV-2 infection within households. People living with HIV (PLHIV) who were not virally supressed were more likely to develop symptomatic illness when infected with SARS-CoV-2, and shed SARS-CoV-2 for longer when compared to HIV-uninfected individuals.

*Implications of all the available evidence:* We found a high rate of SARS-CoV-2 infection in households in a rural community and an urban community in South Africa, with the majority of infections being asymptomatic in individuals of all ages. Asymptomatic individuals transmitted SARS-CoV-2 at similar levels to symptomatic individuals suggesting that interventions targeting symptomatic individuals such as symptom-based testing and contact tracing of individuals tested because they report symptoms may have a limited impact as control measures. Increased household transmission of Beta and Delta variants, likely contributed to recurrent waves of COVID-19, with >60% of individuals infected by the end of follow-up. Higher attack rates, reinfection and acquisition in adolescents and prolonged SARS-CoV-2 shedding in PLHIV who were not virally suppressed suggests that prioritised vaccination of individuals in these groups could impact community transmission.

## Introduction

Many low- and middle-income countries have experienced large numbers of hospitalisations and deaths related to COVID-19. However, reported levels of illness are not always proportional to the high levels of infection implied by serologic studies, suggesting reduced access to laboratory testing, or differences in transmission or susceptibility to developing SARS-CoV-2-related illness in these settings.^1,2,3^ Few detailed SARS-CoV-2 cohort studies are available from middle and low income settings, where vaccination rates remain suboptimal and immunity comes primarily from natural infections, as is the case in South Africa. South Africa has a relatively young population with <5% of the population aged >65 years.^4^ In 2017, the national HIV prevalence among individuals of all ages was 14%, with 7,9 million people living with HIV.^5^

Following the initial detection of SARS-CoV-2 in South Africa in March 2020, a hard lockdown was implemented with restrictions on international travel, school closures, halting of non-essential business and confining people to their homes. Subsequent to this hard lockdown there was a progressive relaxation of restrictions, beginning on 1 May 2020.^6^ South Africa experienced three SARS-CoV-2 waves through August 2021; the first wave peaking in August 2020, the second, associated with the emergence of the Beta variant of SARS-CoV-2, peaking in January 2021 and the third associated with the Delta variant peaking in June 2021.^7^ SARS-CoV-2 restrictions were increased moderately including school closures around the peak of the second and third waves and subsequently relaxed when case numbers decreased. Both Beta and Delta variants have been shown to escape from immunity from previous infection, be more transmissible and may be associated with more severe disease, although the epidemiological consequences of each of these parameters remain debated.^8,9,10,11^

Studies to quantify the burden of asymptomatic infections, symptomatic fraction, duration of shedding and household transmission of SARS-CoV-2 from asymptomatically infected individuals have mostly been conducted as part of outbreak investigations or in specific settings.^12,13,14,15^ Comprehensive community studies of asymptomatic infection and robust individual-level epidemiologic data about infection with Beta and Delta variants are limited.

In randomly selected households from a rural and an urban community in South Africa, we estimated the cumulative incidence of SARS-CoV-2 infection using serial real-time reverse transcription PCR (rRT-PCR) and serology. We estimated the symptomatic fraction of SARS-CoV-2 infection, the duration of viral RNA shedding and the household cumulative infection risk (HCIR) from symptomatic and asymptomatic index cases of different ages.

## Methods

The Prospective Household study of SARS-CoV-2, Influenza and Respiratory Syncytial virus community burden, Transmission dynamics and viral interaction in South Africa (PHIRST-C) was based on a previously conducted study (PHIRST) at the same sites from 2016-2018.^16,17^ We implemented a prospective household cohort study in a rural and an urban community of South Africa with twice weekly collection of mid-turbinate nasal swabs, symptom, and health-seeking data and serum collection every two months to measure SARS-CoV-2 antibodies (Supplementary figure 1). The study included 58 weeks of follow-up at the rural site (16 July 2020 through 28 August 2021) and 56 weeks at the urban site (27 July 2020 through 28 August 2021) with seven serum collections at each site. Nasal swab collection began prior to the first wave peak in the district where the rural site was located and during the peak of the wave in the urban site.^18^

The rural site in Mpumalanga Province is nested within a health and socio-demographic surveillance system (HDSS) run by the Medical Research Council/University of Witwatersrand Rural Public Health and Health Transitions Research Unit, Agincourt.^19,20^ The urban site, Jouberton Township in Klerksdorp, is located in the North West Province.

We aimed to enroll a total of 1000 individuals of all ages. Assuming an average household size of 5 individuals and loss to follow up of 10%, we planned to enroll approximately 110 households from each site (additional information on the sample size calculation is provided in the supplement). Households were randomly selected, from the HDSS database in the rural site and using Global Positioning System (GPS) coordinates in the urban site. Households with >2 members and where ≥80% of eligible members consented to participate were eligible. Details on household selection, enrolment and data collection are provided in the supplement. In brief, we first approached households previously enrolled in PHIRST, and then prospectively approached new potentially-eligible households using the site-specific sampling frame used for PHIRST until the required number of households were enrolled.

### Data collection

We collected individual baseline data, including demographics, HIV status, and history of underlying illness. Study staff visited participating households twice weekly (Monday-Wednesday and Thursday-Saturday) during July 2020-August 2021 to collect mid-turbinate nasal swabs from participants and information about symptoms, absenteeism, and health system contact. Different symptoms were captured among individuals aged <5 years and ≥5 years (Supplementary table 1). Study staff entered data in the field on tablets using REDCap (Research Electronic Data Capture)^21^ and had refresher training on specimen collection and the identification of respiratory signs and symptoms each week.

Study staff collected blood specimens from participants at enrollment (20 July–17 September 2020, blood draw (BD) 1), and every two months thereafter (21 September–10 October, BD2; 23 November–12 December 2020, BD3; and 25 January–21 February 2021, BD4; 22 March–11 April, BD5; 20 May – 9 June 2021, BD6; 19 July – 5 August 2021, BD7). Serology data from the first 5 blood draws have been published.^22^

### Laboratory methods

Nasal specimens were collected using nasopharyngeal nylon flocked swabs, placed in Universal Transport Medium (UTM) and transported daily on ice packs to the National Institute for Communicable Diseases (NICD) in Johannesburg, South Africa, for testing. Nucleic acids were extracted from 200μl of UTM using the Microlab NIMBUS Instrument (Hamilton, Nevada, USA) with the STARMag Universal Cartridge extraction kit (Seegene Inc., Seoul, Korea) according to manufacturer instructions. Specimens were initially tested for the presence of SARS-CoV-2 nucleic acids by rRT-PCR using the Allplex™ 2019-nCoV kit (Seegene Inc., Seoul, Korea) and a BioRad CFX96 thermal cycler, according to manufacturer instructions. From March 2021, samples were tested using the Allplex™ SARS-CoV-2/FluA/FluB/RSV kit (Seegene Inc., Seoul, Korea). A cycle threshold (C_t_) value of <40 on ≥1 of 3 SARS-CoV-2 PCR targets (E, N and S or RdRp genes) was considered positive. All specimens testing SARS-CoV-2-rRT-PCR-positive were confirmed by repeat testing of a second aliquot, and PCR testing in duplicate. Specimens testing positive on at least one duplicate were considered positive. If a specimen was confirmed positive after repeat testing, the results [C_t_ value and targets testing positive] from the first positive test were included in the analysis. A lower Ct-value on rRT-PCR (using the lowest Ct value for any target during the episode) was used as a proxy for higher RNA viral load.^23^ All confirmed positive samples were tested to identify variants of concern using the Allplex™ SARS-CoV-2 Variants I assay (Seegene Inc., Seoul, Korea). This assay targets the RdRp gene, HV69/70 deletion, N501Y and E484K mutations, thus identifying the B.1.351/P1 (Beta/Gamma) and B.1.1.7 (Alpha) variants. From May 2021 SARS-CoV-2-positive samples were also tested using the Allplex™ Variants II assay (Seegene Inc., Seoul, Korea) which detects the L452R mutation (Delta) and differentiates Beta (K417N) from Gamma (K417T). SARS-CoV-2 sequencing methods are described in the supplement.

Serologic evidence of SARS-CoV-2 infection was determined using the Roche Elecsys® Anti-SARS-CoV-2 assay (Roche Diagnostics, Rotkreuz, Switzerland), using recombinant protein representing the nucleocapsid (N) antigen. The assay was performed on the Cobas e601 instrument, and a cut-off index of ≥1.0 was considered an indication of infection (seropositivity).

HIV status was obtained from patient medical records if a participant reported being HIV-infected, or by nurse-administered rapid HIV test with pre- and post-test counselling for participants with unknown, or self-reported HIV-negative status. Patients newly diagnosed with HIV were referred to the nearest primary health care facility for assessment and initiation of antiretroviral treatment.

Individuals were considered fully vaccinated for SARS-CoV-2 ≥14 days after they had received a single dose of the Johnson and Johnson (J&J) vaccine or two doses of the Pfizer BioNTech (Pfizer) vaccine. They were considered partially vaccinated if they had received any vaccine dose but not meeting the above criteria. Only the J&J and Pfizer vaccines were available during the study period.

### Definitions and statistical analyses

We included individuals with ≥10 completed follow-up visits. We defined a SARS-CoV-2 serology-confirmed infected individual as at least one instance of SARS-CoV-2 antibody seropositivity. We defined a SARS-CoV-2 rRT-PCR-confirmed infection episode as at least one nasal swab rRT-PCR positive for SARS-CoV-2. Infection episode duration was estimated from the first to the last day of SARS-CoV-2 rRT-PCR positivity. Details are provided in the supplement (Supplementary figure 2). An illness episode was defined as an episode with ≥1 symptom reported from one visit before, to one visit after the SARS-CoV-2 infection episode. People living with HIV (PLHIV) were deemed to be significantly immunocompromised if their CD4 T-lymphocyte count was <200 cells/μl, and not HIV virally suppressed if their HIV viral load measured ≥400 copies/ml.^24^

An rRT-PCR-confirmed household cluster was composed of all rRT-PCR-confirmed infection episodes within a household within an interval between the rRT-PCR positive tests of any infection episode pairs of ≤14 days (representing ≤2 mean serial intervals).^25^ Cluster duration was estimated as the interval from the first day of rRT-PCR positivity of the first individual in a cluster to the last day of rRT-PCR positivity of the last individual in that cluster. HCIR was defined as the proportion of household members with subsequent infection following SARS-CoV-2 introduction and estimated by dividing the number of subsequent individuals with confirmed infection within a household cluster following SARS-CoV-2 introduction by the number of susceptible (no evidence of previous infection on rRT-PCR or serology) household members. The primary/index case was defined as the first individual testing positive within a cluster on rRT-PCR. Households with co-primary index cases (two individuals rRT-PCR positive for the first time on the same visit) were excluded from the analysis of HCIR.

The generation interval was calculated as the difference between the date of the first positive rRT-PCR in the index and the secondary infection within an rRT-PCR-confirmed household cluster. Following examination of the distribution of calculated generation intervals, we included all secondary infections with rRT-PCR positivity ≤21 days after the index case onset as potential secondary cases for analysis of factors associated with generation interval. Using these definitions, it was possible for a household to experience >1 cluster of infections. Based on epidemic timing in the two communities, first wave episodes or clusters were defined as having onset before 19 December 2020 at both sites, second wave as having onset before 9 June 2021 in Agincourt and 23 June 2021 in Klerksdorp, and third wave as onset up to 28 August at both sites when intense follow-up ended. A variant was allocated to each episode of infection according to a hierarchical process that accounted for known lineages (i.e., wild-type, Alpha, Beta or Delta variant) within episode or household clusters or occurrence of the episode in wave 1, 2 or 3 (see supplement for details). To assess SARS-CoV-2 reinfection, we defined possible reinfection as >28 to 90 days between rRT-PCR -positive specimens (no sequence/variant data available) or between first seropositive specimen and rRT-PCR -positive specimen; probable reinfection as >90 days between rRT-PCR -positive specimens (no sequence/variant data available) or between first seropositive specimen and rRT-PCR -positive specimen; and confirmed reinfection as distinct Nextstrain clades on sequencing or variant PCR between rRT-PCR-positive specimens meeting the temporal criteria for possible or probable.^26^ The proportion of reinfections was calculated as the number of individuals with re-infection divided by the total number of individuals with evidence of prior infection.

For analyses of symptomatic fraction, infection episode duration, HCIR and generation interval we only included incident episodes defined as those occurring with onset >14 days after the start of follow-up. This was because individuals tested positive at the start of follow-up (n=7 and n=32 at the rural and urban site respectively), and we did not know how long they had been shedding SARS-CoV-2, if they had symptoms previously, or who the index case was.

Proportions were compared using the Chi-squared or Fisher’s exact test. We used Weibull accelerated failure time regression, for the analysis of factors associated with time-to-event outcomes (duration of shedding and generation interval). We used logistic regression for the analysis of factors associated with binary outcomes (symptomatic vs. asymptomatic, HCIR, index case vs other participants, reinfection). We used Poisson regression for the analysis of factors associated with at least one SARS-CoV-2 infection episode (cumulative incidence) including all individuals with evidence of infection on rRT-PCR and/or serology. For all analyses we accounted for within-household clustering using random effects regression models. For each multivariable model, we considered all a-priori likely biologically associated factors with the outcome of interest for which we had available data. We examined factors associated with several different outcomes, therefore the selected predictors varied across models. For analyses by age group, in each analysis we chose as reference the age group with the lowest prevalence of the outcome of interest.

Pairwise interactions were assessed graphically and by inclusion of product terms for all variables remaining in the final multivariable additive model. We conducted all statistical analyses using STATA version 14.1 (Stata Corp LP, College Station, Texas, USA). For each univariate analysis, we used all available case information. P values <0.05 were considered statistically significant.

### Ethics

The PHIRST protocol was approved by the University of Witwatersrand Human Research Ethics Committee (Reference 150808) and amended to include enrollment and testing for COVID-19 on 24 June 2020 and was registered on clinicaltrials.gov on 6 August 2015 and updated on 30 December 2020 (https://clinicaltrials.gov/ct2/show/NCT02519803). Informed consent was obtained from all participants prior to data collection. Participants received grocery store vouchers of USD 3 per visit to compensate for time required for specimen collection and interview.

## Results

We approached 537 households, of which 236 (52%) agreed to participate in the study, and of these 222 (94%) were included in the analysis. Of the 1,251 eligible household members, 1,200 (96%) were included. Reasons for non-inclusion are shown in Supplementary figure 3. Among the 222 households in the analysis, the median number of household members was 5 (interquartile range (IQR) 4-7), median sleeping rooms was 3 (IQR 2-4), and 49% had at least one child aged <5 years, with a higher proportion among households in the rural community (Supplementary table 2). Individuals from the rural community were younger, had a lower level of formal education and were less likely to be employed. Underlying illness was more common in the urban site, but HIV prevalence was similar between sites (14% in the rural and 17% in the urban site, p=0.173). At the end of follow up 5% of individuals were fully vaccinated againast SARS-CoV-2.

At the start of follow up in July 2020, 1% (5/443) and 15% (73/498) of individuals with available data had serologic evidence of previous SARS-CoV-2 infection at the rural and the urban site, respectively. Of 125,088 potential individual follow-up visits, we collected and tested 115,759 (93%) mid-turbinate nasal swabs, of which 1976 (2%) tested positive for SARS-CoV-2 on rRT-PCR (Figure 1 and Supplementary figure 5 and 6). During the study, 90% (200/222) of households had at least one individual testing SARS-CoV-2 positive on rRT-PCR or serology, with an average of 3.7 (range 1-10) infected individuals per infected household.

**Figure 1:**
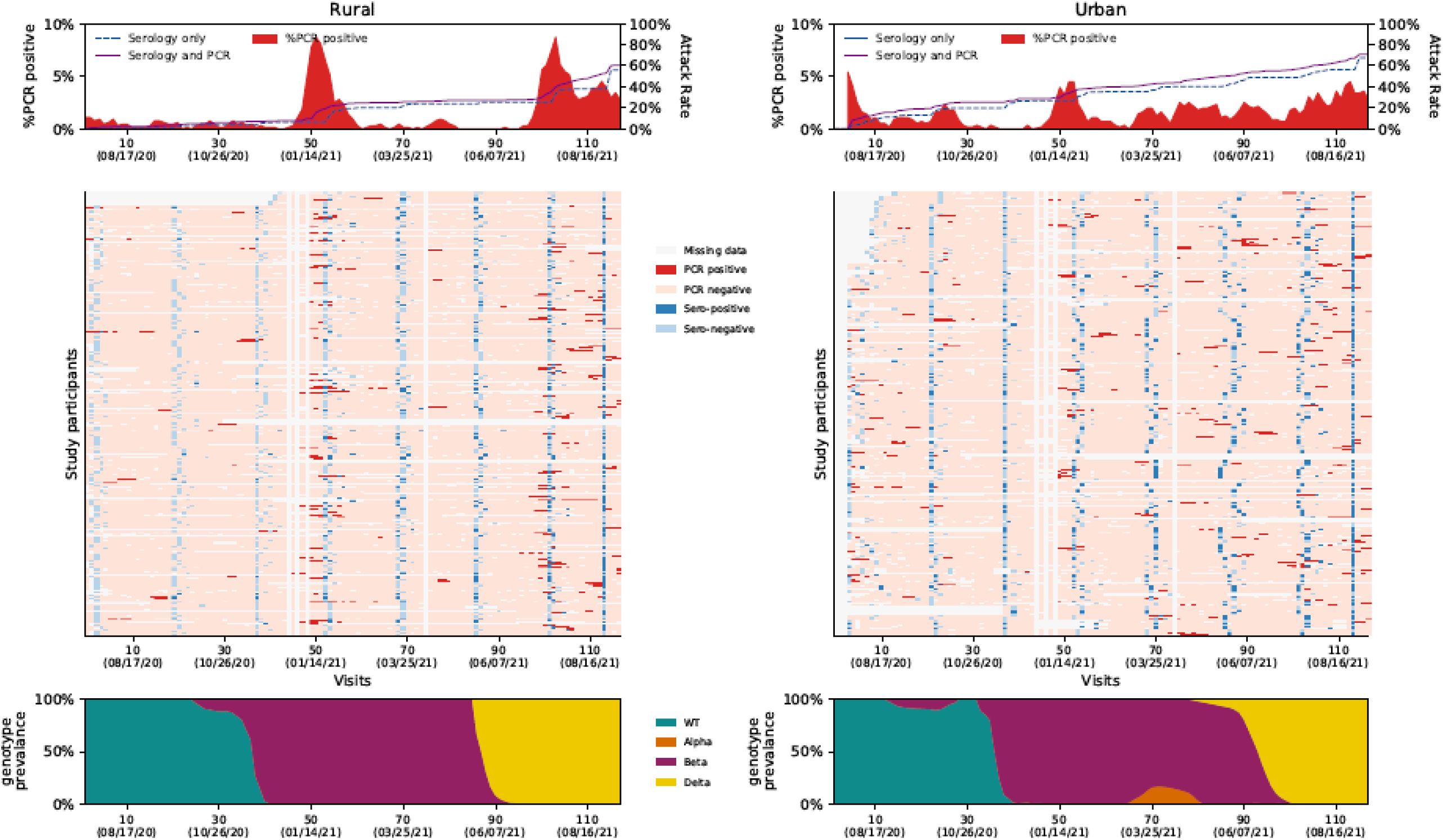
Top panel: Percentage testing real-time reverse transcription polymerase chain reaction (rRT-PCR)-positive per study visit and cumulative percentage with evidence of infection (attack rate) on serology only and on rRT-PCR and serology combined, a rural site and an urban site, South Africa, 2020-2021. Middle panel: Results of serology and rRT-PCR of individuals enrolled in the PHIRST-C study, a rural site and an urban site, South Africa, 2020-2021. Columns are individual follow up visits and rows are individual participants. Individuals within the same household are numbered consecutively (appear below one another). Follow up visits are coloured white if no sample was tested, light pink if the sample tested negative for SARS-CoV-2 and coloured red if the nasopharyngeal swab tested positive for SARS-CoV-2. Cells at the time of serology blood draws are coloured according to the results of serology as follows: light blue - serology negative, dark blue - serology positive. Bottom panel: Percent of rRT-PCR-positive samples typed as wild type (WT) or variant of concern (Alpha, Beta or Delta) by follow-up visit (10 visit moving average used for smoothing).

During the follow-up period, 62% (749/1200) of individuals experienced at least one episode of SARS-CoV-2 infection on rRT-PCR and/or serology and 12% (87/749) experienced a repeat infection, including one individual who experienced two repeat infections. Of 88 repeat infection episodes, 10 (11%) were classified as possible, 21 (24%) as probable and 57 (65%) as confirmed (Figure 2 and supplement). The highest proportion infected was in the 13-18 years age group (Supplementary figure 4). Repeat infection was more common in individuals aged 13-18 years and in the urban site (Supplementary Table 3).

**Figure 2:**
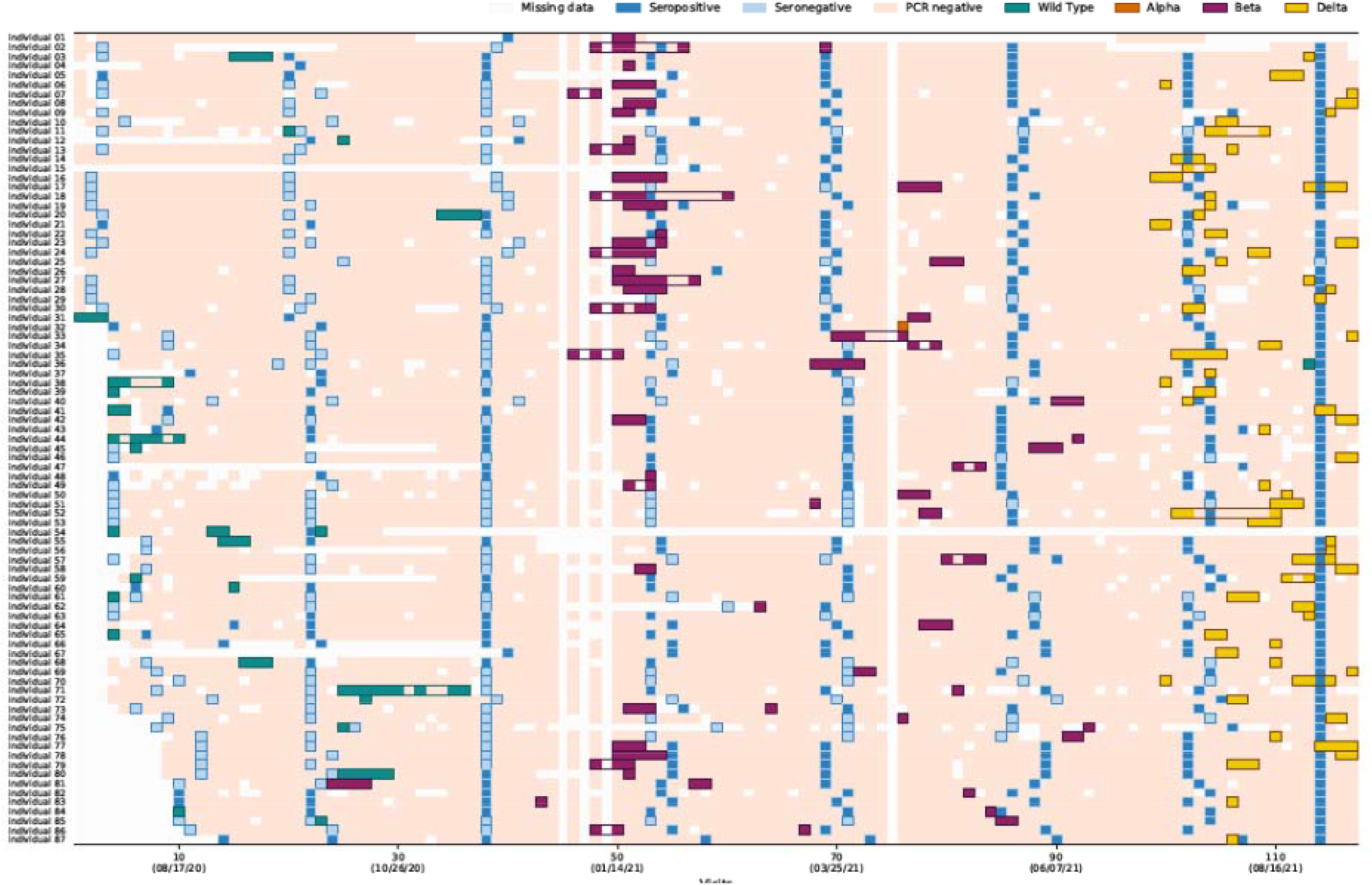
Timing of results of serology and real-time reverse transcription polymerase chain reaction (rRT-PCR)-of 87 individuals with definite, probable or possible reinfections, a rural site and an urban site, South Africa, 2020-2021 Columns are individual follow up visits and rows are individual participants. Follow up visits are coloured white if no sample was tested, light pink if the sample tested negative for SARS-CoV-2. PCR-positive follow up visits are coloured different colours according to infecting variant. Infection episodes are outlined in corresponding colours. Within an episode some visits may test negative or be missed, these are coloured light pink or white. Cells at the time of serology blood draws are coloured according to the results of serology as follows: light blue serology negative, dark blue serology positive.

Among 294 individuals with a positive rRT-PCR during follow-up who had a negative serology preceding the episode and available serology data >14 days after the start of the episode, 267 (91%) seroconverted after the episode. Failure to develop a serologic response was more common in individuals aged <5, 19-39 and 40-59 years (compared to 5-12 years) and episodes with a single rRT-PCR positive swab and with Ct value>30 (Supplementary table 4). Among 447 individuals who were seronegative at baseline and subsequently became seropositive, 404 (90%) had evidence of a rRT-PCR-confirmed infection.

On multivariable analysis, factors associated with SARS-CoV-2 cumulative incidence were age 5-12 and 13-18 years vs <5 years, residing in the urban community vs rural community, and being overweight vs normal weight (Table 1).

**Table 1:**
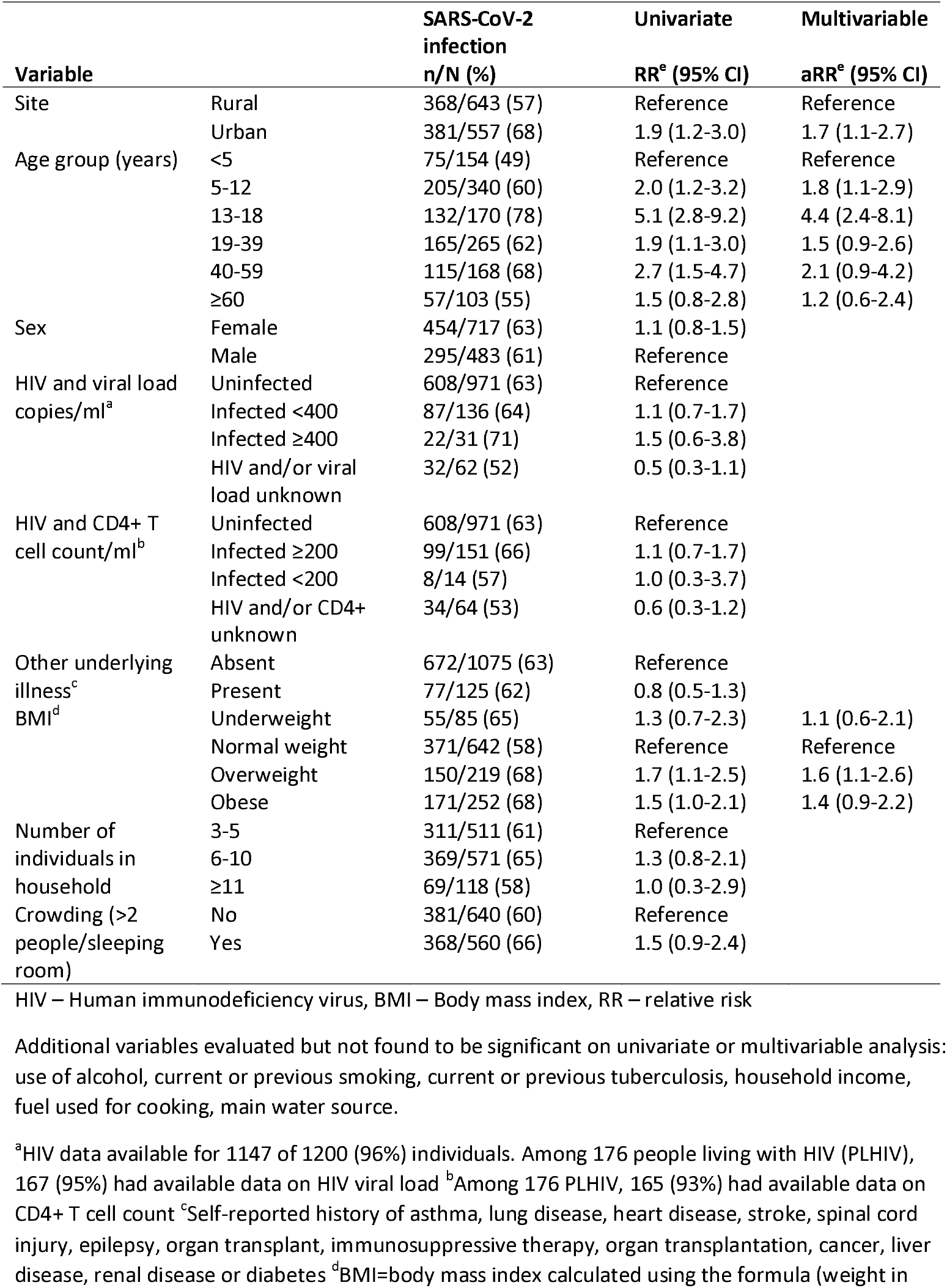
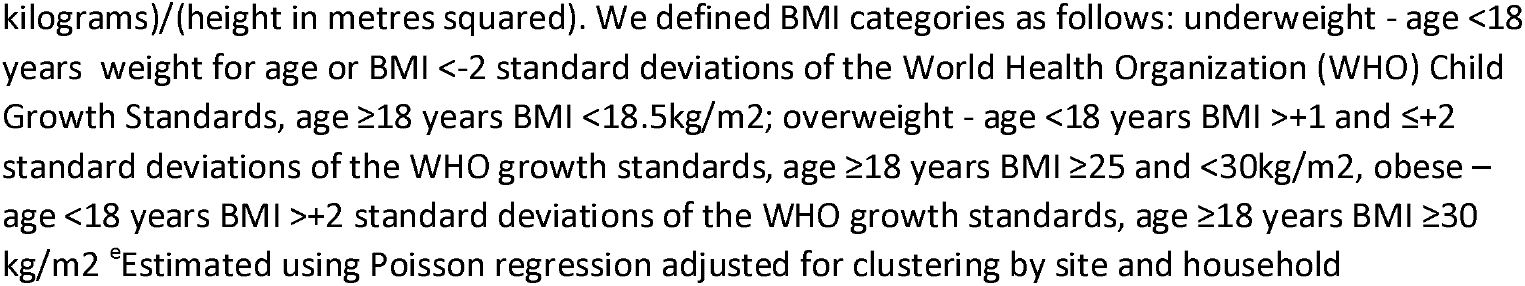
Factors associated with cumulative incidence of severe acute respiratory syndrome coronavirus 2 (SARS-CoV-2) infection on real-time reverse transcription polymerase chain reaction (rRT-PCR) and/or serology in a rural and an urban community, South Africa, 2020-2021

Of 718 rRT-PCR-confirmed episodes, 17% (n=124) occurred in the first wave, 55% (n=270) in the second wave and 45% (n=324) in the third wave (Supplementary figure 5 and 6). Proportionately more children and adolescents were infected with successive waves with 43% (53/124) of cases in wave one aged <19 years compared to 49% (133/270) in the second and 66% (213/324) in the third wave respectively (p<0.001). Of 662 rRT-PCR-confirmed episodes, that occurred >14 days after the start of follow-up, 15% (97/662) of individuals reported ≥1 symptom (details in supplement). Among 97 symptomatic individuals, 6 (6%) attended an outpatient clinic, 9 (9%) were hospitalised and 2 (2%) died (infection fatality ratio 0.3% (95% confidence interval (CI) 0.03%-1%)). Among 20 symptomatic individuals who were employed or attended school, 35% (n=7) reported absenteeism. In multivariable analysis symptoms were more common in individuals aged ≥19 years compared to <5 years, in PLHIV with HIV viral load ≥400 copies/ml, in obese individuals and in episodes with minimum Ct value <30 and caused by Delta variant (Table 2). Positive serology before the episode was associated with decreased symptoms.

**Table 2:**
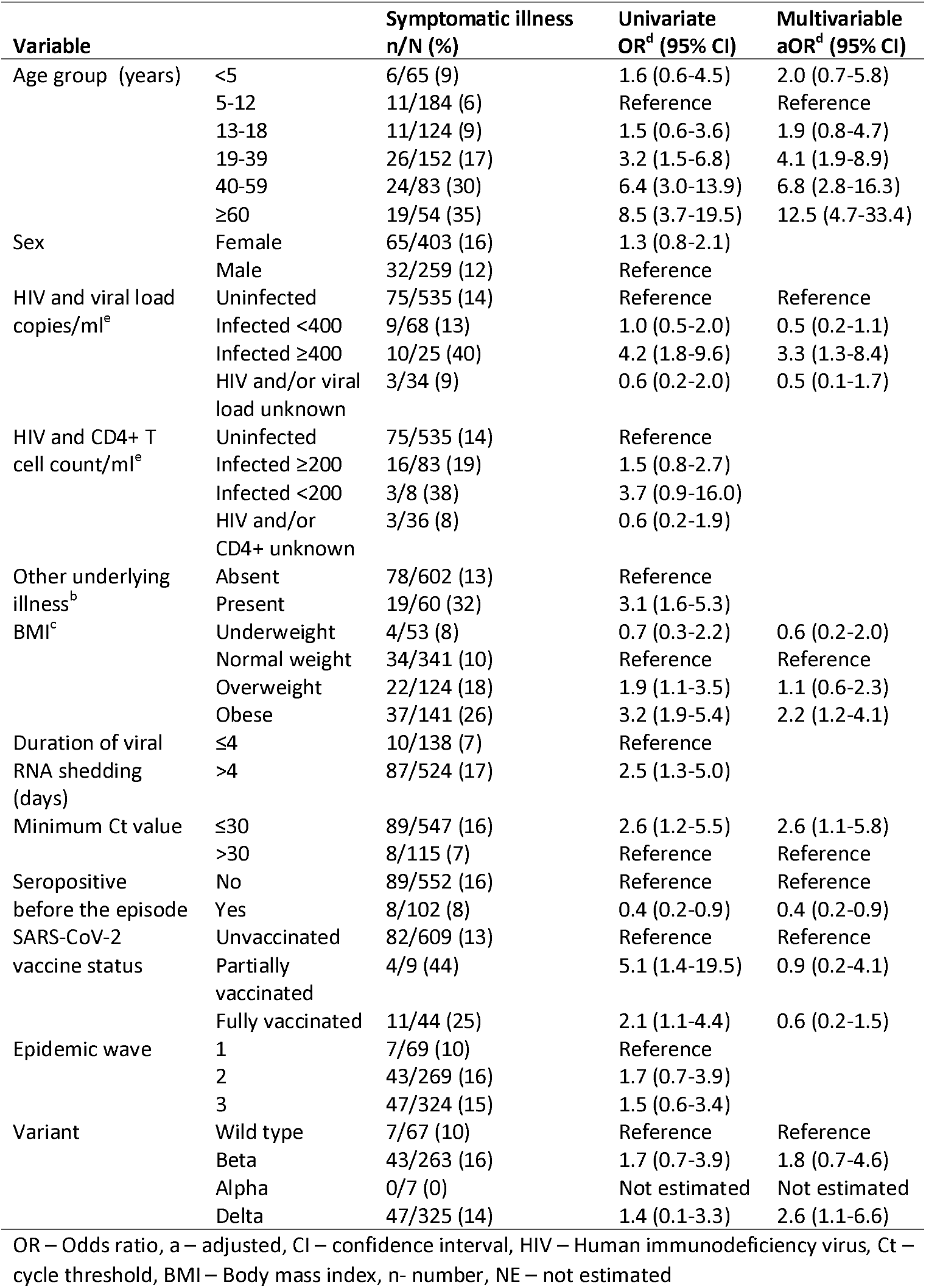
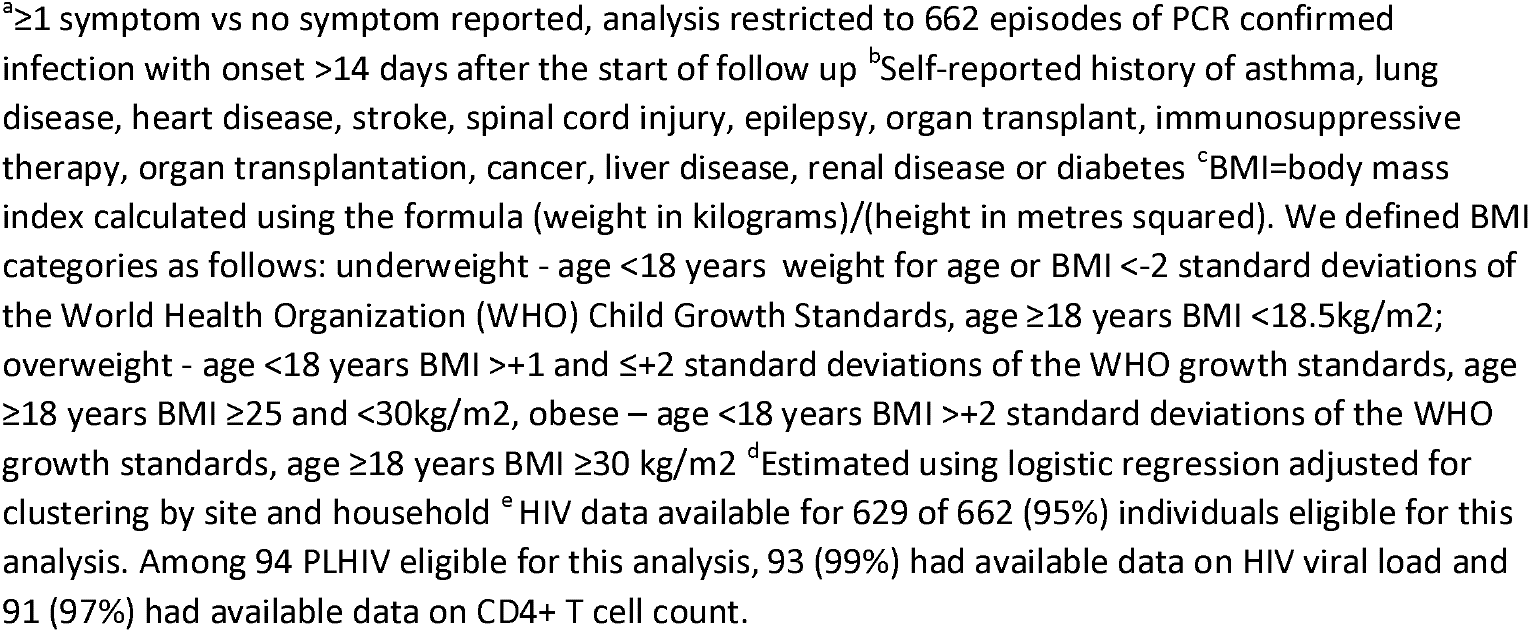
Factors associated with symptomatic illness^a^ among real-time reverse transcription polymerase chain reaction (rRT-PCR)-confirmed severe acute respiratory syndrome coronavirus 2 (SARS-CoV-2)-infected individuals in a rural and an urban community, South Africa, 2020-2021

The mean duration of rRT-PCR positivity was 11.6 days (median 11 days, range 4-137 days) and 21% (138/662) of episodes were rRT-PCR positive at only one visit. On multivariable analysis, symptomatic individuals, PLHIV with HIV viral load ≥400 copies/ml and individuals with minimum Ct value <30 shed SARS-CoV-2 RNA for longer (Table 3). Positive serology before the episode was associated with decreased duration of shedding.

**Table 3:**
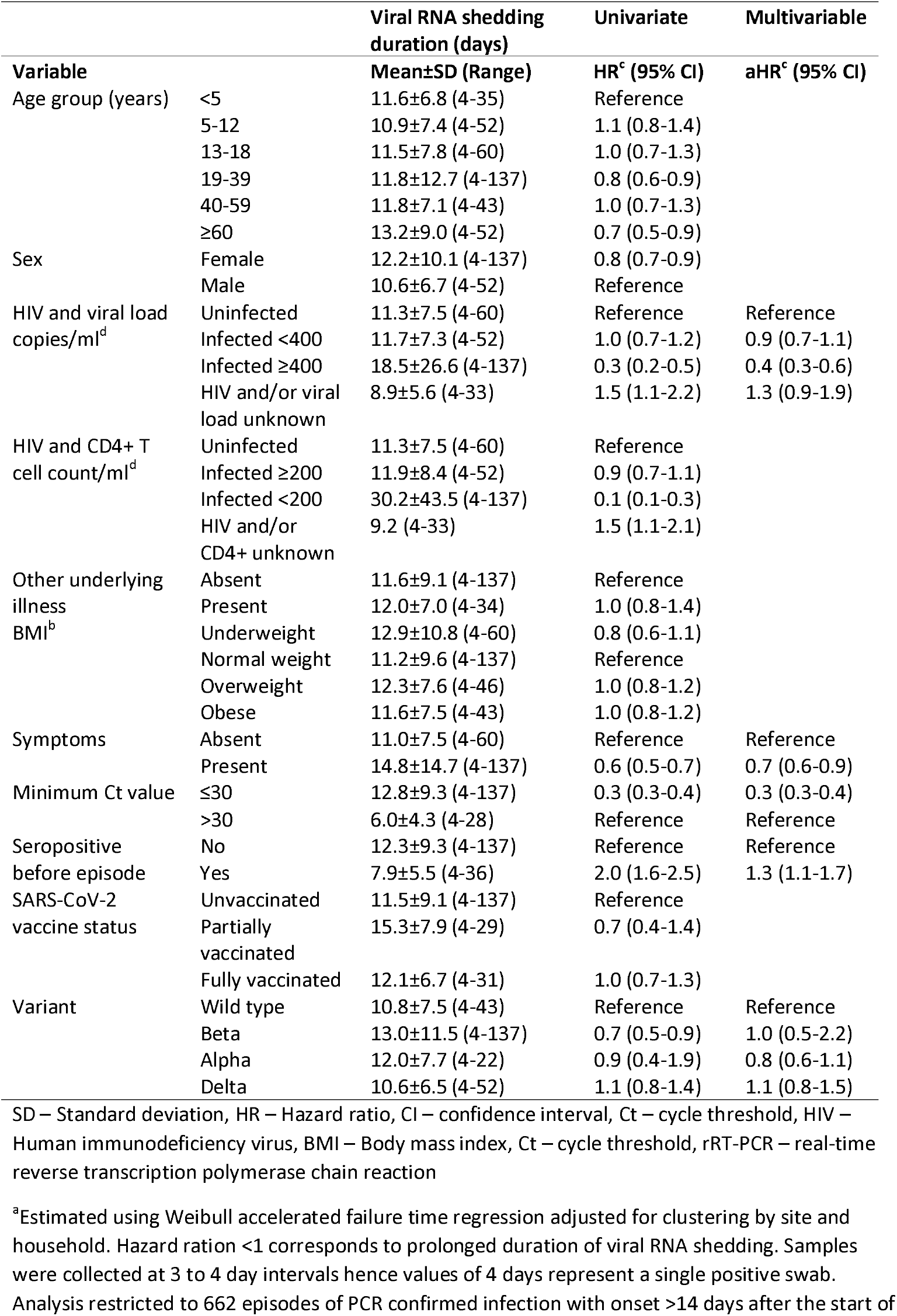
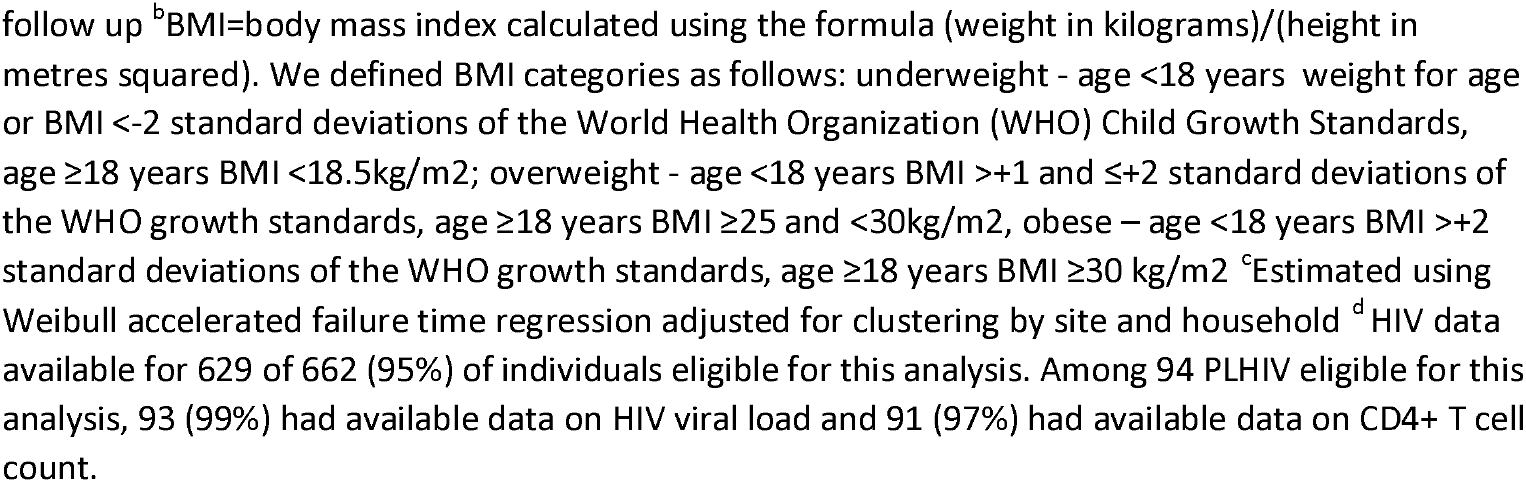
Factors associated with duration of severe acute respiratory syndrome coronavirus 2 (SARS-CoV-2) viral RNA positivity in a rural and an urban community, South Africa, 2020-2021^a^

Among 195 households with at least one SARS-CoV-2 infection cluster detected on rRT-PCR, 72 (37%) had one cluster, 83 (43%) had 2 clusters, 31 (16%) had 3 clusters and 9 (5%) had >3 clusters (total of 369 clusters). The average cluster duration among 336 clusters starting >14 days after the start of follow up was 15.6 days (range 4-137 days). We included 184 clusters starting >14 days after the start of follow up and with a single index case from 103 households for analysis of HCIR. In this subset of households the HCIR was 25% (213/856, 95% CI 22%-28%). HCIR was 19% (21/110) for symptomatic and 25% (195/775) for asymptomatic index cases (p=0.165) (Table 4). On multivariable analysis, low Ct value (proxy for high viral RNA load) of the index case, index case female sex, index case age 40-59 years and household contact age between 5 and 18 years, urban community and infection with Beta and Delta variant were associated with increased HCIR.

**Table 4:**
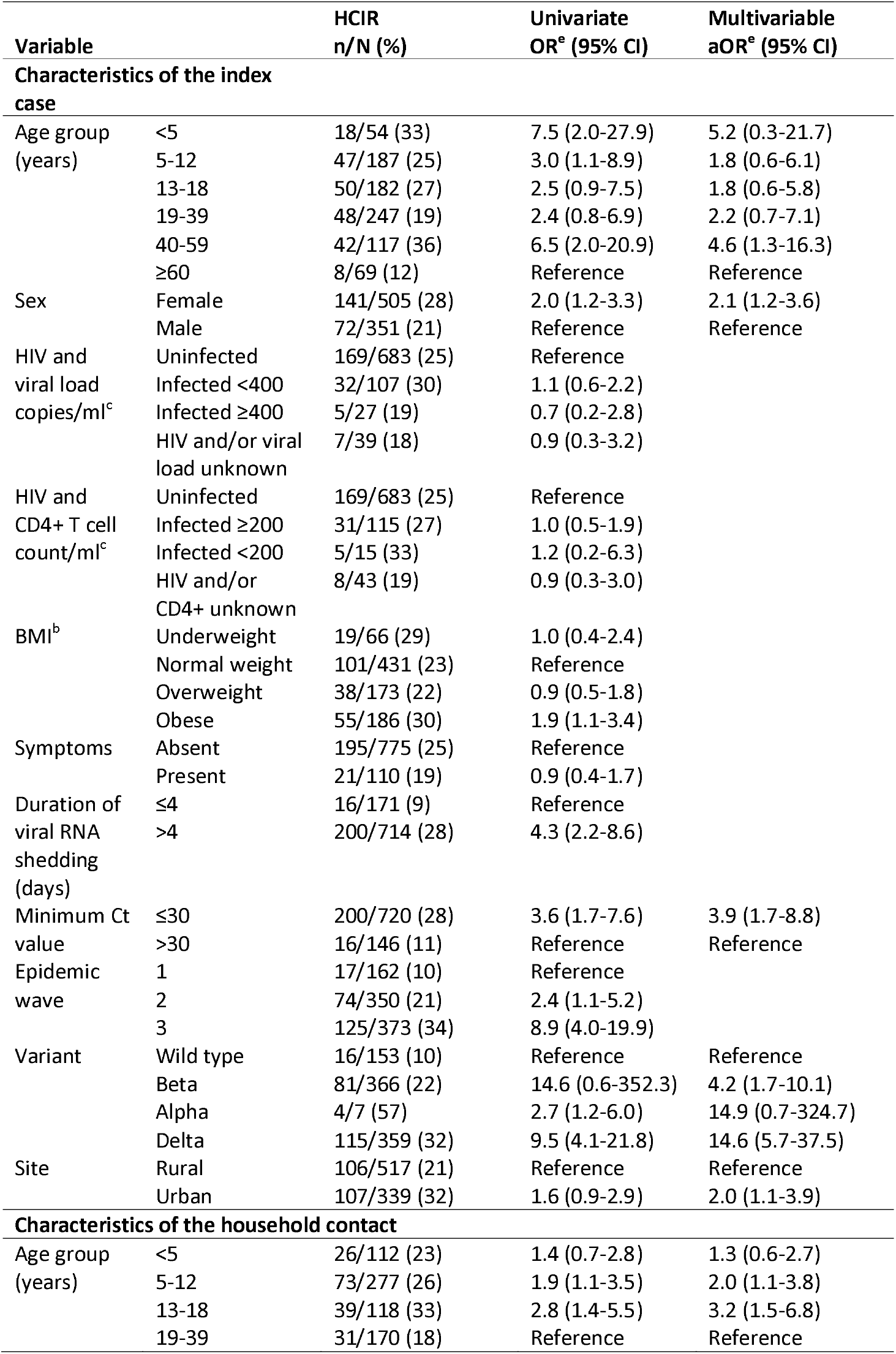
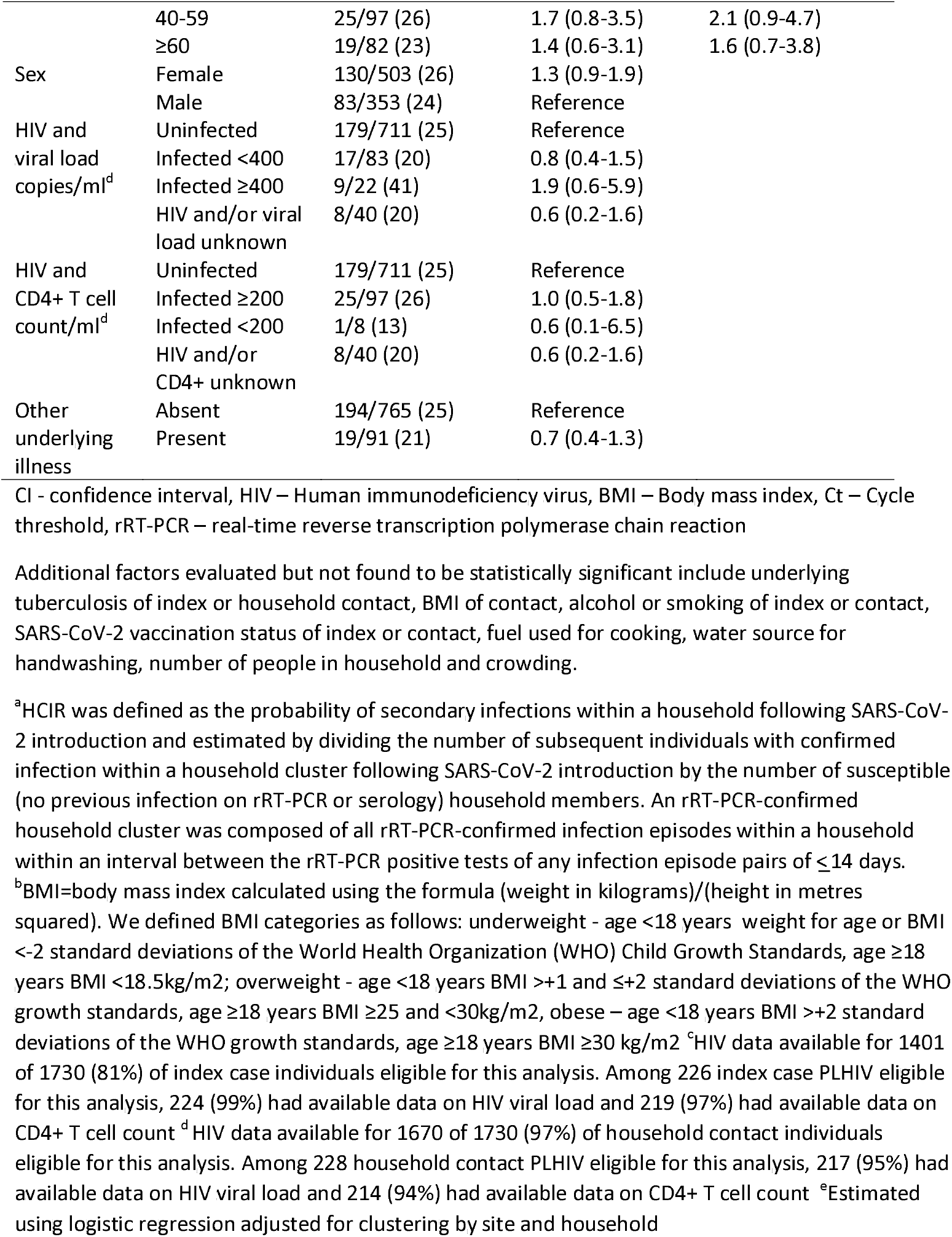
Factors associated with household cumulative infection risk (HCIR)^a^ in a rural and an urban community, South Africa, 2020-2021

The mean generation interval was 7.5 days (range 2-21 days) (Supplementary figure 7). On multivariable analysis, generation interval was shorter if the index case was symptomatic and longer if the index case shed for longer or was infected with Alpha, Beta or Delta variant (Supplementary table 4).

## Discussion

Using intensive systematic repeated sampling among household cohorts in two largely unvaccinated South African communities, we found that by 18 August 2021 almost two-thirds of individuals had been infected with SARS-CoV-2. Approximately 12% of individuals experienced at least one repeat episode of infection within 13 months of follow up. Only 15% of infections were associated with symptoms; of these, 9% were hospitalised and 2% died. SARS-CoV-2 Delta variant infections were more likely to be symptomatic compared to non-variant infection. In households with at least one SARS-CoV-2 infection, SARS-CoV-2 was transmitted to 25% of household contacts irrespective of symptoms in the index case. Index and household contact age, infecting SARS-CoV-2 variant and index case RNA viral load were the main predictors of onward transmission. Household transmission was increased fourfold with the Beta variant and fifteenfold with the Delta variant.

Previous studies have generated wide-ranging estimates of the proportion of SARS-CoV-2 infections which are asymptomatic. A recent systematic review found that 20% (95% CI 3%-67%) of SARS-CoV-2 infections remained asymptomatic throughout infection and that transmission was lower from asymptomatic individuals.^27^ We found that 85% of infections were asymptomatic despite active twice-a-week symptom evaluation. Three US-based cohort studies implementing weekly PCR testing for SARS-CoV-2, irrespective of symptoms, found estimates of 31% asymptomatic in a community-based study including children, 35% in pregnant women and 11% among healthcare workers, lower than estimates in our study. ^28–30^ The lower symptomatic fraction in these studies could be because twice weekly-swabbing allows identification of more transient symptomatic illness or because there could be truly lower symptomatic fraction in the populations included in our study. Compared to other studies, the population included in our study was relatively young with only 9% of individuals aged ≥60 years, reflecting the general South African population.^4^ Despite the low overall symptomatic fraction, symptomatic fraction increased with age, in line with prior studies.^15,27^

A systematic review and meta -analysis of 87 household transmission studies of SARS-CoV-2 reported an estimated secondary attack rate of 18.9% (95% CI 16.2-22.0)^31^ slightly lower than our estimate of 25%. The review also reported that household secondary attack rates were higher from symptomatic index cases, that adults were more likely than children to acquire infection and that sex was not associated with transmission. Previous studies have found that viral load and duration of SARS-CoV-2 shedding are lower among mild and asymptomatically infected individuals compared to individuals with severe illness.^32^ While we did not find that symptom profile was associated with risk of infection or transmission, we did find that higher index case viral RNA load (estimated through the proxy of cycle threshold value) was associated with more frequent transmission. This is similar to a study from Catalonia where viral load was carefully quantified in adult transmitters.^33^ We also found females were more likely to transmit SARS-CoV-2, potentially reflecting closer contact with other household members in this population. Several studies have demonstrated increased transmissibility of the Beta and Delta SARS-CoV-2 variants.^9,34^ We found that household transmission of SARS-CoV-2 was approximately 4 times higher with Beta and 15 times higher with Delta variant compared to non-variant infections. It is possible that participants changed their behaviour once informed that they were infected with SARS-CoV-2, however data on behaviour following a SARS-CoV-2 diagnosis were not available.

Symptomatic fraction was lowest in children and adolescents aged <18 years. While attack rates were lower in children aged <5 years, they were highest in adolescents aged 13-18 years. We also found that indviduals aged 13-18 years were more likely to acquire infection within the household and more likely to experience reinfection with SARS-CoV-2. Previous studies have found lower attack rates and symptomatic fraction in children (although relatively increased in age group 13-18 years, similar to our finding) and also that children are less likely to transmit SARS-CoV-2 and have reduced susceptibility to infection.^35–37,38,39^ The Delta variant has been associated with higher attack rates in children and adolescents in South Africa and elsewhere. In several countries, this has in part been attributed to a shift in the age distribution of cases as vaccination expanded to adult age groups.^40^ In South Africa, where vaccination remains low, infection rates were higher among adults in the first two waves, potentially contributing to the higher attack rate in children and adolescents with Delta variant. Differences in circulating variants over time and geography may have contributed to differences in the contribution of adolescents to transmission in previous studies.^41^ In addition, most previous studies did not include systematic longitudinal rRT-PCR testing irrespective of symptoms in children, adolescents and adults, potentially biasing against detection of minimally symptomatic infections in children.

Previous studies have found that PLHIV are more likely to be hospitalised and that hospitalised PLHIV without HIV viral suppression shed SARS-CoV-2 at a high viral load for longer, but data from community settings are lacking.^42–44^ We found that PLHIV who were not virally suppressed were more likely to develop symptomatic illness when infected with SARS-CoV-2, and shed SARS-CoV-2 for longer when compared to HIV-uninfected individuals, potentially contributing to the evolution of novel variants of SARS-CoV-2.

Our study had several strengths. Participating households were randomly sampled from a rural and an urban South African community and followed up for 13 months through the second half of the first wave and the second and third waves of SARS-CoV-2 infection in South Africa. Participating individuals were sampled twice weekly, irrespective of symptoms, allowing for more accurate ascertainment of the burden of SARS-CoV-2 infections as well as the symptomatic fraction and transmission from asymptomatic individuals. Our unique study design combining frequent rRT-PCR and serological testing allowed for more complete ascertainment of infection burden.

Our study also had several limitations. We included two communities and households with >2 members, potentially limiting generalisability of study findings. The finding of different attack rates in the two communities suggests substantial heterogeneity in disease transmission in different geographic areas. Participants were sampled using mid-turbinate nasal swabs because of potential SARS-CoV-2 transmission risk with a collection of more sensitive nasopharyngeal swab specimens. This could have led to some missed infections. However, the strong association between rRT-PCR- and serology-confirmed infection in individuals with both specimen types available suggests that the majority of infections were detected. Repeated questioning on symptoms twice weekly may be associated with participant fatigue and under-reporting. Participants may have been informed of their SARS-CoV-2 infection status before developing symptoms, potentially affecting reporting. We implemented several measures to reduce this potential bias including weekly retraining of field workers on symptom collection and regular field supervisory visits to evaluate data collection and symptom recording. A study of influenza infection in the same population with similar study design found that 56% of individuals infected with influenza were symptomatic, suggesting the robustness of our data.^17^ We did not quantify viral RNA load but instead this was inferred using Ct values as proxy.

In conclusion, we found a high rate of SARS-CoV-2 infection in households in a rural and an urban South African community with the majority of infections being asymptomatic in individuals of all ages. Individuals aged 13-18 years had the highest attack rates and were more likely to acquire infection. The household cumulative infection risk was 25% and did not differ by presence of symptoms in index case but was higher in households where the index case had a higher RNA viral load and with Beta and Delta variant infection. Asymptomatic individuals transmitted SARS-CoV-2 at similar levels as symptomatic individuals suggesting that interventions targeting symptomatic individuals, such as promotion of community testing and contact tracing of individuals tested because they report symptoms, may have limited impact in this setting.

## Data Availability

The investigators welcome enquiries about possible collaborations and requests for access to the data set. Data will be shared after approval of a proposal and with a signed data access agreement. Investigators interested in more details about this study, or in accessing these resources, should contact the principle investigator, Prof Cheryl Cohen, at NICD.

## Acknowledgements

The authors would like to thank all the individuals who kindly agreed to participate in the study as well as the many field and laboratory staff who worked tirelessly to implement the study. The MRC/Wits Rural Public Health and Health Transitions Research Unit and Agincourt Health and Socio-Demographic Surveillance System, a node of the South African Population Research Infrastructure Network (SAPRIN), is supported by the Department of Science and Innovation, the University of the Witwatersrand, the Medical Research Council, South Africa, and the Wellcome Trust, UK (grants 058893/Z/99/A; 069683/Z/02/Z; 085477/Z/08/Z; 085477/B/08/Z).

## Author’s contributions

Conception and design of study: CC, JK, AvG, MLM, NW, JM, NAM, KK, LL, ST

Data collection and laboratory processing: CC, JK, AvG, MLM, NW, JNB, JM, MdP, MC, AB, NAM, KK, ST, LL, FW, JdT, FXG, FSD, TMK, ST

Analysis and interpretation: CC, JK, AvG, MLM, NW, JNB, JM, MdP, MC, AB, NAM, KK, STo, LL, FW, JdT, FXG, FSD, TMK, KS, CV, ST

CC, JK, and ST accessed and verified the underlying data. CC and ST drafted the Article. All authors critically reviewed the Article. All authors had access to all the data reported in the study.

## Disclaimer

The findings and conclusions in this paper are those of the authors and do not necessarily represent the official position of the funding agencies.

## Ethics

The PHIRST-C protocol was approved by the University of Witwatersrand Human Research Ethics Committee (Reference 150808) and the U.S. Centers for Disease Control and Prevention’s Institutional Review Board relied on the local review (#6840). The protocol was registered on clinicaltrials.gov on 6 August 2015 and updated on 30 December 2020 (https://clinicaltrials.gov/ct2/show/NCT02519803).

## Potential conflicts of interest

CC has received grant support from Sanofi Pasteur, Advanced Vaccine Initiative, and payment of travel costs from Parexel. AvG has received grant support from Sanofi Pasteur, Pfizer related to pneumococcal vaccine, CDC and the Bill & Melinda Gates Foundation. NW report grants from Sanofi Pasteur and the Bill & Melinda Gates Foundation. NAM has received a grant to his institution from Pfizer to conduct research in patients with pneumonia and from Roche to collect specimens to assess a novel TB assay.

The full study protocol can be found on the National Institute for Communicable Diseases Website at the following link: https://www.nicd.ac.za/centres/centre-for-respiratory-disease-and-meningitis/

