## Supplement for "SARS-CoV-2 incidence, transmission and reinfection in a rural and an urban setting: results of the PHIRST-C cohort study, South Africa, 2020-2021"

**Supplemental methods**

**Household selection and participant enrolment**

We randomly selected households using different methods in each site. In Agincourt, for the original A Prospective Household cohort study of Influenza, Respiratory Syncytial virus and other respiratory pathogens community burden and Transmission dynamics in South Africa (PHIRST) study, in 2017 and 2018 we selected two villages out of 29 within the Health and Demographic Surveillance Site (HDSS) according to convenience. Within these villages, households with >2 members were randomly selected. We only included households with >2 household members as smaller households could provide limited information on household transmission. For the current study, we approached all houses participating in the initial PHIRST study in 2017 and 2018 and if sufficient numbers were not enrolled we approached additional households from the same villages using the same sampling frame and ensuring similar total numbers of included households from all four villages. We did not approach houses participating in PHIRST in 2016 at the rural site because logistically it was more feasible to conduct the study in four (rather than six) villages.

In Jouberton Township, for the original PHIRST study, a list of 450 random global positioning system (GPS) coordinates were generated in the study area using Google Earth as previously described.^1^ Study staff navigated to the coordinates and selected the nearest house within 30 meters of the location. If there was no dwelling within 30m the coordinates were discarded. We approached all houses participating in the initial PHIRST study from 2016 through 2018 and if sufficient numbers were not enrolled we approached additional households from the same township using the same sampling frame until the desired sample size was reached. We approached houses participating in PHIRST from all three years (2016-2018) at the urban site because logistically this was feasible due to the compact size and random distribution of enrolled households.

Household size was similar for included houses and general community at the rural site (number of household members=4, interquartile range 1-20 in study vs n=4 interquartile range 1-14 in community) and at the urban site (number of household members=5, interquartile range 3-14 in study vs n=4 interquartile range 3-5 in community)^2^.

**Sample size**

We aimed to study infection and reinfection in a total of 1000 individuals. Assuming an average household size of 5 individuals and loss to follow up of 10%, we planned to enrol approximately 110 households from each site. Assuming a constant community attack rate of 60% across 5 selected age groups (based on an estimated *R_0_* value of 2.5^3^) for a 95% confidence interval, a 10% desired absolute precision and a 1.3 design (household cluster) effect, 120 individuals were needed in each age stratum. The expected age distribution in the target cohort was 16% for individuals aged <5 years (160 expected individuals in the cohort), 41% for individuals aged 5-18 years (410 expected individuals in the cohort), 26% for individuals aged 19-44 years (260 expected individuals in the cohort), 12% for individuals aged 45-64 (120 expected individuals in the cohort) and 5% for individuals aged ≥65 years (50 expected individuals in the cohort).

We assumed 30% symptomatic fraction among individuals infected with SARS-CoV-2.^4^ We assumed 20% severe cases and a 2% mortality rate among all symptomatic individuals. The sample sizes to estimate the above parameters for a 95% confidence interval and a 10% desired absolute precision are as follows: (i) 30% symptomatic fraction among infected individuals: 81 infected individuals; (ii) 20% severe cases among symptomatic individuals: 62 symptomatic individuals severe; (iii) 2% mortality among symptomatic individuals: 8 symptomatic individuals died. A total of 270 individuals with SARS-CoV-2 infection (i.e., 81 symptomatic individuals/0.3 symptomatic fraction; community attack rate: 27.0%) will be needed to estimate the above-mentioned parameters with the desired level of precision.

**Staff and participant safety**

A number of measures were introduced to minimise risks to the study team and potentially to participants due to the circulation of SARS-CoV-2. Front-line staff including study nurses were trained in infection control procedures including proper hand hygiene and the correct use of surgical face masks, not only to minimize their own risk of infection when in close contact with patients during home visits and elsewhere, but also to minimize the risk of the nurses acting as a vector of infection between household members or between households. Staff were required to wear gloves for collection of patient specimens. All staff were offered influenza vaccination prior to commencing the study, and again before the influenza season in 2021. Staff were properly trained in collection of blood specimens and disposal of sharps. Staff were requested to stay home from work if they had symptoms of respiratory illness or if they had been in contact with a confirmed of suspected SARS-CoV-2 case without appropriate personal protective equipment. Retraining of all safety procedures including field implementation evaluation was performed weekly. Additional measures implemented included use of dedicated staff transport adhering to government regulations on occupancy numbers, provision of scrubs, headcover, aprons and surgical masks for fieldwork, consistent physical distancing for all staff, frequent disinfection of all equipment between households, use of hand sanitiser and daily temperature checks. All interviews were conducted from a distance of at least 2 metres from the interviewer outside of the house if possible and with no physical contact between participants and study staff member. The study convened a safety advisory board (SAB) to advise investigators on the safety procedures in the study including the appropriate PPE, safety procedures for interaction with infected household members, other workplace safety concerns. The SAB members were independent of the study and constituted clinical, microbiology and infection prevention and control experts. A community representative was included from each site.

Individuals testing positive for SARS-CoV-2 were notified through the NMC (Notifiable Medical Conditions) system and rapidly communicated to the provincial Departments of Health (Mpumalanga and North West provinces) and contact tracing was performed by local public health authorities. Participants were advised to isolate if they tested positive for SARS-CoV-2 or if they screened positive for COVID-like symptoms whilst awaiting results. As per National guidelines, isolation was advised for 10 days from the onset of symptoms or from the date of testing in the case of an asymptomatic case.

Nurses counselled participants to isolate in a separate room of the house, to limit contact with other household members, to wear a mask when leaving their isolation room to use the bathroom, to wipe down items used in the common space (such as taps or toilet), to not share food, and to sanitise regularly. The household was also advised to ensure good ventilation within the house (opening windows and doors).

Due to the nature of the rural and urban environment, the adoption of these measures was adversely affected by numerous factors. Those without enough rooms attempted to lower transmission risk by minimising the number of people sleeping in the same room, limiting the time spent in the same room as the index case, having the index case wear a mask whilst with household members, and opening windows. Where a child was the index case, the mother or primary caregiver would stay with the child to care for them. Many found it difficult to limit the movement of, and interactions with, SARS-CoV-2 positive children especially their interactions with other children in the household. Many participants were asymptomatic during their SARS-CoV-2 infection and this combined may have negatively impacted adherence to recommended measures. The average turn-around time for samples from collection to returning result to the participant was 5 days for symptomatic individuals and 10-14 days for asymptomatic individuals leading to delays in implementation of recommended measures.

A nasopharyngeal swab was collected from all field workers on a weekly basis and tested for SARS-CoV-2 to document possible infection transmission to and by fieldworkers. Any positive test was considered an adverse event and were reported to the study SAB.

**Participant enrolment**

In the enrolment visit, field staff first confirmed that the household contained >2 members and then requested permission from the head of household to inform members about the study. After a minimum of three failed attempts, if the head of household was unavailable or a minor, the household was excluded. Study staff requested informed consent to participate in the study from all household members aged ≥18 years, assent from children aged 7 to 17 years, and consent from a parent or guardian for children younger than 18 years. If a household withdrew their participation while in the first 6 months of the study follow up, they were replaced with a newly recruited household.

**Data collection**

Symptom data collection and temperature measurement were performed at each follow up visit. Symptoms collected included fever (self-reported or measured tympanic temperature ≥38◦C), cough, difficulty breathing, sore throat, nasal congestion, vomiting, diarrhoea, abdominal pain or loss of smell or taste, muscle aches, fatigue, headache and confusion. Symptom data collected differed by age group to account for age-specific differences in clinical presentation and difficulty collecting information on subjective symptoms such as muscle aches or headache from young children. Symptoms collected in each age group are detailed in supplementary table 1. Symptom data for children was consistently collected from the individual identified as primary caregiver at the time of enrolment. For older children and adolescents, self-reported symptom data were also accepted. Field workers received refresher training on different aspects of study implementation including data collection, specimen collection and use of online databases at least weekly. Field workers were encouraged to observe participants for visible symptoms such as cough and prompt for symptoms if not reported. Measured tympanic temperature was recorded at each study visit and either reported or measured fever was included as presence of fever. Site supervisors conducted regular (at least monthly) supervisory visits to assess study implementation in the field and external (teams from outside the study sites) supervisory visits were conducted quarterly. During the two weeks in December over the Christmas and New Year holidays, and again at Easter home visits for collection of mid-turbinate nasal swabs were conducted only once a week instead of twice-weekly.

Written vaccination history was obtained for all children aged <5 years from patient-held immunisation records and, if needed, vaccination records at health facilities. Primary caregivers giving a history of the child never being vaccinated were recorded as unvaccinated.

Vaccination against SARS-CoV-2 commenced in South Africa in February 2021, initially among health care workers using the Johnson & Johnson (J&J) vaccine as part of a Phase 3 study, followed by the initiation of national vaccination programme including the Pfizer BioNTech and J&J vaccines in individuals ≥60 years of age from May 2021, sequentially moving to 50-59 year olds and 35-49 year olds by the end of follow up.

Household income was evaluated through self-reporting by the head of household rounded to the nearest rand. For households where income varied from month to month, the head of household was asked to provide the average monthly household income over the previous 12 months.

Infants were defined as HIV exposed but uninfected if they were HIV-uninfected but the mother was living with HIV. For people living with HIV (PLHIV), samples were collected for determination of CD4 T-lymphocyte count and HIV quantitative viral load testing at diagnosis or enrolment and current use of antiretroviral therapy was documented.

**Laboratory methods**

All samples testing rRT-PCR-positive were confirmed by re-extraction and rRT-PCR testing of a second aliquot in duplicate. Specimens testing positive on ≥2 of three rRT-PCRs were considered positive. If a sample was confirmed positive the Ct-value and the number of targets positive from the first positive test were used for the analysis. For the first 46 visits this testing was done retrospectively after 1 to 6 months of storage at -70 degrees centigrade. Following this, confirmatory resting was performed in real time following identification of a rRT-PCR-positive sample.

We confirmed presence of human DNA in samples through testing for the RNaseP gene on a random 10% of samples weekly until 17 March 2021 and subsequently tested all samples for presence of RNaseP. Of 13581 samples tested for RNaseP, human DNA was detected in 13122 (97%).

For sequencing of SARS-CoV-2, we used AmpliSeq for SARS-CoV-2 (Illumina), which is based on the ARTIC SARS-CoV-2 sequencing protocol (https://www.protocols.io/view/illumina-nextera-dna-flex-library-construction-and-bhjgj4jw), on the Ion Torrent Genexus platform. Genomes were assembled using the Exatype SARS-CoV-2 pipeline (https://sars-cov-2.exatype.com/), which includes de-duplicating sequenced reads data, base-calling, de-multiplexing, removal of amplicon primer sequences prior to variant calling, mapping to a reference, polishing and finally consensus sequence generation. Clade and lineage assignments were made using the online Nextclade (https://clades.nextstrain.org/) and Pangolin (https://pangolin.cog-uk.io/) applications, which also enable identification of known variants of concern as well as novel mutations. We used in-house R codes and AliView (https://ormbunkar.se/aliview/) to polish sequences and display analyses. In addition we used a Galaxy SARS-CoV-2 pipeline to analyse minority variant analysis.

**Definitions and statistical analyses**

A variant was allocated to each episode of infection according to the following hierarchical process:

1. At least one sample within the identified episode of infection with confirmed rRT-PCR wild-type or variant result: wild-type or variant assigned to the entire episode.
2. No samples within the identified episode of infection with confirmed rRT-PCR wild-type or variant result, but the episode is within a household cluster with at least one known episode of infection with confirmed rRT-PCR wild-type or variant result: cluster wild-type or variant assigned to the episode.
3. No samples within the identified episode of infection with confirmed rRT-PCR wild-type or variant result, and the episode is not within a household cluster with at least one known episode of infection with confirmed rRT-PCR wild-type or variant result: wild-type or variant assigned to the episode based on wave as a proxy for lianeage circulation (i.e., wave 1: wild-type; wave 2: Beta variant; wave 3: Delta variant).

For the analysis of factors associated with SARS-CoV-2 infection on PCR or serology we assessed overdispersion starting with a negative binomial model and retained a simpler Poisson model as there was no statistically significant overdispersion detected*.*

**Ethics**

The U.S. Centers for Disease Control and Prevention’s Institutional Review Board relied on the local review (#6840).

**Disclaimer**

The findings and conclusions in this report are those of the author(s) and do not necessarily represent the official position of the funding agencies or the Centers for Disease Control and Prevention.

**Supplementary results**

**Characteristics of reinfections**

There were 87 reinfections identified during follow up (10 possible, 20 probable and 57 confirmed), 31 at the rural site and 56 at the urban site. Median age was 24 years (range 1-71 years), 58 (67%) were female, 16 (20%) of 81 with available data were PLHIV of whom 15 had data on CD4+ T cell count and HIV viral load and 1 had CD4+T cell count <200 cells/mm^3^ and 4 had ≥1000 HIV viral copies per ml plasma. Eight (9%) individuals with reinfection had non-HIV underlying illness.

**Symptoms and illness severity**

Of 662 episodes, that occurred >14 days after the start of follow-up, 15% (97/662) of individuals reported ≥1 symptom, with 5% (30/662) reporting 1 symptom and 10% (67/662) reporting ≥2 symptoms. The most commonly reported symptoms were cough (12%, 78/662), runny nose (7%, 43/662) and headache (4%, 24/598). Among 662 episodes of infection, only 3% (17/662) reported difficulty breathing, 2% (14/662) reported fever and 2% (12/662) reported fever with cough. Other symptoms reported were diarrhoea 1% (4/662), vomiting 1% (5/662), abdominal pain 1% (4/598), sore throat 3% (18/598), body pains 2% (11/598), fatigue 3% (20/598), loss of smell or taste 2% (10/598) and confusion <1% (2/598).

There were 9 hospitalised individuals, all at the urban site. Median age of hospitalised individuals was 62.2 years (range 38-79 years) and 4 (4/9) were female. Of the nine hospitalised individuals one (1/7) was PLHIV, 4 (4/9) had non-HIV underlying illness, 4 (4/9) were overweight and 4 (4/9) were obese, none (0/7) had a smoking history and 5 (5/9) reported previous alcohol use.

**Characteristics of index cases vs other cases within clusters**

When compared to non-index cases, index cases within household clusters were more likely to be aged 13-59 years compared to <5 years (Supplementary table 5).

Supplementary table 1: Symptom data collected among children aged <5 years and individuals aged ≥5 years in the PHIRST-C study

| **Symptom** | **Age group** |  |
| --- | --- | --- |
|  | **<5 years** | **≥5 years** |
| Measured fever | Collected | Collected |
| Reported fever | Collected | Collected |
| Cough | Collected | Collected |
| Shortness of breath | Not collected | Collected |
| Difficulty breathing or chest in drawing | Collected | Not collected |
| Sore throat | Not collected | Collected |
| Nasal congestion or runny nose | Collected | Collected |
| Lost sense of smell or taste | Not collected | Collected |
| Feeding poorly or had little appetite | Collected | Not collected |
| Vomiting | Collected | Collected |
| Diarrhoea (3 or more loose stools in 24 hours) | Collected | Collected |
| Abdominal pain | Not collected | Collected |
| Muscle aches | Not collected | Collected |
| Fatigue for one or more days | Not collected | Collected |
| Headache | Not collected | Collected |
| Been confused or unable to respond to questions | Not collected | Collected |
| Irritable or inconsolable | Collected | Not collected |
| Lethargic (unable to walk, sit, feed) | Collected | Not collected |
| Other symptoms (free text field) | Collected | Collected |

Supplementary table 2: Baseline characteristics of households and individuals included in a Prospective Household cohort study of Influenza, Respiratory Syncytial virus and other respiratory pathogens community burden and Transmission dynamics in South Africa – COVID version (PHIRST-C) in a rural and an urban community, South Africa, 2020-2021

| **Characteristic** | **Overall**  **n (%) or median (IQR)** | **Rural**  **n (%) or median (IQR)** | **Urban**  **n (%) or median (IQR)** | **OR (95% CI)^k^**  **Urban vs. Rural** |
| --- | --- | --- | --- | --- |
| **Household level characteristics** | **N=222** | **N=114** | **N=108** |  |
| Number of household members  3-5  6-10  >10 | 131 (59)  82 (37)  9 (4) | 62 (54)  48 (42)  4 (4) | 69 (60)  34 (31)  5 (5) | Reference  0.6 (0.4-1.1)  1.1 (0.3-4.3) |
| Median number of household members | 5 (4-7) | 5 (4-7) | 5 (4-6) |  |
| Number of rooms  1-4  5-9  ≥10 | 76 (34)  138 (62)  8 (4) | 38 (33)  69 (61)  7 (6) | 38 (35)  69 (64)  1 (1) | Reference  0.9 (0.5-1.5)  0.1 (0.0-1.1) |
| Median number of rooms | 5 (4-7) | 5 (3-8) | 5 (4-6) |  |
| Number of rooms for sleeping  1-2  2-4  >4 | 93 (42)  106 (48)  23 (10) | 37 (32)  57 (50)  20 (18) | 56 (52)  49 (45)  3 (3) | Reference  0.5 (0.3-0.9)  0.1 (0.1-0.3) |
| Median number of rooms for sleeping | 3 (2-4) | 3 (2-4) | 2 (2-3) |  |
| Crowding (>2 people/sleeping room) | 83 (37) | 38 (33) | 45 (42) | 1.4 (0.8-2.5) |
| Child aged <5 years in house | 109 (49) | 67 (59) | 42 (39) | 0.4 (0.2-0.8) |
| Household member smokes indoors | 59 (27) | 13 (11) | 46 (43) | 5.8 (2.9-11.5) |
| Main water source tap inside (vs tap outside) | 135 (61) | 58 (51) | 77 (71) | 2.4 (1.4-4.2) |
| Handwashing place with water in house | 207 (93) | 103 (90) | 104 (96) | 2.8 (0.9-9.0) |
| Main fuel for cooking  Electricity  Wood  Paraffin/gas/other | 141 (64)  79 (36)  2 (1) | 35 (31)  79 (69)  0 (0) | 106 (98)  0 (0)  2 (2) | Reference  0.0 (0.0-0.1)  1.7 (0.1-34.5) |
| Monthly household income^a^  ≤R800 (<USD54)  R801-R1600 (USD55-108)  R1601-R3200 (USD109-116)  R3201-R6400 (USD117-232)  R6401-R12800 (USD233-464)  >R12800 (>USD464) | 12 (6)  38 (18)  90 (43)  54 (26)  11 (5)  3 (1) | 8 (7)  15 (13)  48 (42)  33 (29)  9 (8)  1 (1) | 4 (4)  23 (24)  42 (45)  21 (22)  2 (2)  2 (2) | Reference  3.1 (0.8-12.0)  1.8 (0.5-6.0)  1.3 (0.3-4.8)  0.4 (0.1-3.1)  4.0 (0.3-58.6) |
| **Individual level characteristics** | **N=1200** | **N=643** | **N=557** |  |
| Age group (years)  <5  5-12  13-18  19-39  40-59  ≥60 | 154 (13)  340 (28)  170 (14)  265 (22)  168 (14)  103 (9) | 99 (15)  211 (33)  88 (14)  131 (20)  68 (11)  46 (7) | 55 (10)  129 (23)  82 (15)  134 (24)  100 (18)  57 (10) | Reference  1.1 (0.7-1.6)  1.7 (1.1-2.6)  1.8 (1.2-2.8)  2.6 (1.7-4.2)  2.2 (1.3-3.7) |
| Female sex | 717 (60) | 409 (64) | 308 (55) | 0.7 (0.6-0.9) |
| Level of education^b^  No schooling  Primary schooling  Some secondary  Secondary completed  Post-secondary | 59 (11)  89 (17)  230 (43)  142 (27)  15 (3) | 35 (14)  44 (18)  83 (34)  80 (33)  3 (1) | 24 (8)  45 (16)  147 (51)  62 (21)  12 (4) | Reference  1.5 (0.8-2.9)  2.6 (1.4-4.6)  1.1 (0.6-2.1)  5.8 (1.5-22.9) |
| Employment^b^  Unemployed  Employed  Student  Pensioner | 352 (66)  27 (5)  109 (20)  47 (9) | 171 (70)  10 (4)  38 (16)  26 (11) | 181 (62)  17 (6)  71 (24)  21 (7) | Reference  1.6 (0.7-3.6)  1.8 (1.1-2.8)  0.8 (0.4-1.4) |
| Reported alcohol use^c^ | 196 (30) | 37 (12) | 159 (47) | 6.3 (4.2-9.5) |
| Reported current cigarette smoking^c^ | 124 (19) | 12 (4) | 112 (33) | 11.9 (6.4-22.1) |
| HIV status^d^  Uninfected  Infected  Unknown | 971 (85)  176 (15)  53 | 520 (86)  84 (14)  39 | 451 (83)  92 (17)  14 | Reference  1.3 (0.9-1.7) |
| HIV viral load^e^  ≥400 copies/ml | 31 (19) | 11 (14) | 20 (21) | 1.9 (0.8-4.2) |
| CD4+ T cell count^f^  <200/ml | 14 (8) | 5 (6) | 9 (11) | 1.8 (0.6-5.5) |
| Previous tuberculosis | 40 (3) | 11 (2) | 29(5) | 3.1 (1.6-6.4) |
| Current tuberculosis | 5 (<1) | 1 (<1) | 4 (1) | 4.6 (0.5-41.6) |
| Other underlying illness^g^ | 125 (10) | 42 (7) | 83 (15) | 2.5 (1.7-3.7) |
| Influenza vaccination 2020 | 22 (2) | 18 (3) | 4 (1) | 0.3 (0.1-0.7) |
| Influenza vaccination 2021 | 7 (1) | 6 (1) | 1 (<1) |  |
| Fully vaccinated against SARS-CoV-2 vaccine by end of follow up^l^ | 57 (5) | 23 (4) | 34 (6) | 1.8 (1.0-3.0) |
| Pneumococcal vaccine up to date for age^j^  Yes  No  No data | 109 (92)  9 (8)  36 | 73 (97)  2 (3)  24 | 36 (84)  7 (16)  12 | 7.3 (1.4-37.1)  Reference |
| DTaP-IPV/Hib vaccine up to date for age^i^  Yes  No  No data | 113 (96)  5 (4)  36 | 75 (100)  0 (0)  24 | 38 (88)  5 (12)  12 | 22.2 (1.2-411.5)  Reference |

HIV – Human immunodeficiency virus, DTaP-IPV/Hib – Diphtheria, tetanus, acellular pertussis, inactivated polio, *Haemophilus influenzae* type B vaccine, IQR – interquartile range, OR – odds ratio, CI – confidence interval, NE – not estimated, n - number, USD – United States Dollar. Penalized logistic regression used for cells with zero values.

^a^Data available for 208 households, 114 rural and 94 urban ^b^Individuals aged >18 years with available data N=535, 245 at rural site and 290 at urban site ^c^Individuals aged ≥15 years N=643, 303 at rural site and 340 at urban site ^d^% and p value among individuals with known status ^e^Among 176 PLHIV, 166 (94%) reported currently receiving antiretroviral treatment (ART), of 165 PLHIV with data on CD4+ T cell count, 151 (92%) were >200 /ml, of 166 individuals with viral load data, 136 (82%) had <400 copies/ml ^g^Self-reported history of asthma, lung disease, heart disease, stroke, spinal cord injury, epilepsy, organ transplant, immunosuppressive therapy, organ transplantation, cancer, liver disease, renal disease or diabetes ^j^Individuals aged <5 years N=154, 99 at rural site and 55 at urban site, 118 with available vaccination data, 75 at the rural site and 43 at the urban site ^k^Estimated using logistic regression adjusted for clustering by site and household ^l^ Of 57 individuals who were fully vaccinated by the end of follow up, 23 received a single dose of the Johnson and Johnson vaccine and 34 received 2 doses of Pfizer vaccine and 47 (82%) were vaccinated from June through September. An additional 58 individuals received the first dose of Pfizer vaccine during the follow up period.

Supplementary Table 3: Factors associated with repeat infection with severe acute respiratory syndrome coronavirus 2 (SARS-CoV-2) infection on real-time reverse transcription polymerase chain reaction (rRT-PCR) and/or serology among 749 individuals with at least one infection in a rural and an urban community, South Africa, 2020-2021

|  |  | **SARS-CoV-2 reinfection** | **Univariate** | **Multivariable** |
| --- | --- | --- | --- | --- |
| **Variable** |  | **n/N (%)** | **RR^e^ (95% CI)** | **aRR^e^ (95% CI)** |
| Site | Rural  Urban | 32/368 (9)  56/381 (15) | Reference  1.9 (1.1-3.0) | Reference  1.8 (1.1-3.0) |
| Age group (years) | <5  5-12  13-18  19-39  40-59  ≥60 | 4/75 (5)  18/205 (9)  24/132 (18)  25/165 (15)  12/115 (10)  5/57 ((9) | Reference  1.6 (0.5-5.1)  4.0 (1.3-12.5)  3.0 (0.9-9.3)  2.0 (0.6-6.7)  1.5 (0.4-6.3) | Reference  1.7 (0.5-5.1)  3.8 (1.3-11.8)  2.9 (0.9-8.5)  1.8 (0.5-5.9)  1.4 (0.4-5.7) |
| Sex | Female  Male | 58/454 (13)  30/295 (10) | 1.4 (0.8-2.2)  Reference |  |
| HIV and viral load copies/ml^a^ | Uninfected  Infected <400  Infected ≥400  HIV and/or viral load unknown | 66/608 (11)  11/87 (13)  4/22 (18)  7/32 (22) | Reference  1.2 (0.6-2.5)  1.9 (0.6-6.1)  2.5 (0.9-6.4) |  |
| HIV and CD4+ T cell count/ml^b^ | Uninfected  Infected ≥200  Infected <200  HIV and/or CD4+ unknown | 66/608 (11)  14/99 (14)  1/8 (13)  7/34 (21) | Reference  1.4 (0.7-2.6)  1.3 (0.1-11.9)  2.3 (0.9-5.7) |  |
| Other underlying illness^c^ | Absent  Present | 80/672 (12)  8/77 (10) | Reference  0.8 (0.4-1.9) |  |
| BMI^d^ | Underweight  Normal weight  Overweight  Obese | 7/55 (13)  37/371 (10)  19/150 (13)  24/171 (14) | 1.3 (0.5-3.2)  Reference  1.2 (0.7-2.3)  1.4 (0.8-2.6) |  |
| Number of individuals in household | 3-5  6-10  ≥11 | 39/311 (13)  42/369 (11)  7/69 (10) | Reference  0.9 (0.5-1.5)  0.7 (0.3-1.9) |  |
| Crowding (>2 people/sleeping room) | No  Yes | 45/381 (12)  43/368 (12) | Reference  1.0 (0.6-1.6) |  |

HIV – Human immunodeficiency virus, BMI – Body mass index, RR – relative risk

Additional variables evaluated but not found to be significant on univariate or multivariable analysis: use of alcohol, current or previous smoking, current or previous tuberculosis, household income, fuel used for cooking, main water source, SARS-CoV-2 vaccination

^a^HIV data available for 719/749 (96%) of individuals. Among 111 PLHIV, 109 (98%) had available data on HIV viral load ^b^Among 111 PLHIV, 107 (98%) had available data on CD4+ T cell count ^c^Self-reported history of asthma, lung disease, heart disease, stroke, spinal cord injury, epilepsy, organ transplant, immunosuppressive therapy, organ transplantation, cancer, liver disease, renal disease or diabetes ^d^BMI=body mass index calculated using the formula (weight in kilograms)/(height in metres squared). We defined BMI categories as follows: underweight - age <18 years weight for age or BMI <-2 standard deviations of the World Health Organization (WHO) Child Growth Standards, age ≥18 years BMI <18.5kg/m2; overweight - age <18 years BMI >+1 and ≤+2 standard deviations of the WHO growth standards, age ≥18 years BMI ≥25 and <30kg/m2, obese – age <18 years BMI >+2 standard deviations of the WHO growth standards, age ≥18 years BMI ≥30 kg/m2 ^e^Estimated using logistic regression adjusted for clustering by site and household

Supplementary table 4: Characteristics associated with failure to develop a detectable serologic response for individuals who had a negative serology result preceding a rRT-PCR confirmed infection with SARS-CoV-2 in a rural and an urban site, South Africa, 2020-2021^a^

|  |  | **No serologic response detected** | **Univariate** | **Multivariable** |
| --- | --- | --- | --- | --- |
| **Variable** |  | **n/N (%)** | **RR (95% CI)** | **aRR (95% CI)** |
| Age group (years) | <5  5-12  13-18  19-39  40-59  ≥60 | 6/30 (20)  3/67 (4)  3/49 (6)  8/75 (11)  5/45 (11)  2/28 (7) | 4.9 (1.2-19.4)  Reference  1.4 (0.3-6.4)  2.3 (0.6-8.6)  2.5 (0.6-10.1)  1.7 (0.3-9.4) | 18.8 (3.1-114.8)  Reference  3.7 (0.6-22.9)  6.0 (1.2-29.0)  7.0 (1.2-40.6)  2.5 (0.3-19.1) |
| Sex | Female  Male | 20/187 (11)  7/107 (7) | 1.6 (0.7-3.9)  Reference |  |
| HIV and viral load copies/ml^c^ | Uninfected  Infected <400  Infected ≥400  HIV and/or viral load unknown | 23/237 (10)  2/33 (6)  2/16 (13)  0/8 (0) | Reference  0.7 (0.2-2.8)  1.6 (0.4-6.4)  Not estimated |  |
| HIV and CD4+ T cell count/ml^c^ | Uninfected  Infected ≥200  Infected <200  HIV and/or CD4+ unknown | 23/237 (10)  3/43 (7)  1/4 (25)  0/10 (0) | Reference  0.8 (0.2-2.5)  3.9 (0.5-27.6)  Not estimated |  |
| Other underlying illness^b^ | No  Yes | 23/262 (9)  4/32 (13) | Reference  1.6 (0.5-4.7) |  |
| Symptoms | Absent  Present | 24/245 (10)  3/49 (6) | Reference  0.7 (0.2-2.3) |  |
| Duration of viral RNA shedding (days) | ≤4  >4 | 21/49 (43)  6/245 (2) | Reference  0.1 (0.1-0.1) | Reference  0.1 (0.0-0.2) |
| Minimum Ct value | ≤30  >30 | 10/253 (4)  17/41 (41) | 0.1 (0.0-0.1)  Reference | 0.2 (0.1-0.1)  Reference |

^a^Estimated using logistic regression adjusted for clustering by site and household ^b^Self-reported history of asthma, lung disease, heart disease, stroke, spinal cord injury, epilepsy, organ transplant, immunosuppressive therapy, organ transplantation, cancer, liver disease, renal disease or diabetes ^c^HIV data available for 286 (97%) of 294 included individuals, of 49 HIV-infected individuals viral load data available for 49 (100%) and CD4+ T cell count data available for 47 (96%)

Supplementary table4: Factors associated with generation interval among 198 individuals with interval ≤21 days at a rural and an urban site, South Africa, 2020-2021^a^

|  |  | **Interval (days)** | **Univariate** | **Multivariable** |
| --- | --- | --- | --- | --- |
| **Variable** |  | **Mean±SD (Range)** | **HR** | **aHR** |
| **Characteristics of the index case** | |  |  |  |
| Age group (years) | <5  5-12  13-18  19-39  40-59  ≥60 | 7.0±4.8 (2-18)  6.6±4.2 (2-20)  9.3±5.7 (2-21)  6.6±4.0 (2-19)  7.7±4.4 (2-20)  6.5±2.4 (3-10) | 0.7 (0.1-1.7)  0.8 (0.4-1.8)  0.5 (0.2-0.9)  0.9 (0.4-1.8)  0.7 (0.3-1.4)  Reference |  |
| Sex | Female  Male | 7.1±4.4 (2-21)  8.5±5.1 (2-21) | 1.4 (1.0-1.8)  Reference |  |
| HIV and viral load copies/ml | Uninfected  Infected <400  Infected ≥400  HIV and/or viral load unknown | 7.6±4.9 (2-21)  7.4±4.2 (3-16)  7.0±5.0 (2-12)  6.1±2.3 (3-10) | Reference  1.1 (0.8-1.6)  1.2 (0.4-3.7)  1.7 (0.8-3.7) |  |
| HIV and CD4+ T cell count/ml | Uninfected  Infected ≥200  Infected <200  HIV and/or CD4+ unknown | 7.6±4.9 (2-21)  6.7±3.8 (2-15)  13.7±2.1 (12-16)  6.3±2.1 (3-10) | Reference  1.3 (0.9-1.9)  0.5 (0.1-1.4)  1.7 (0.8-3.5) |  |
| Symptoms | Absent  Present | 7.7±4.8 (2-21)  5.6±3.6 (3-15) | Reference  1.7 (1.1-2.7) | Reference  1.7 (1.1-2.7) |
| Minimum Ct value | ≤30  >30 | 7.6±4.8 (2-21)  7.0±3.1 (3-14) | 8 (0.5-1.4)  Reference |  |
| Duration of shedding (days) | ≤4  >4 | 5.4±3.4 (2-14)  7.7±4.8 (2-21) | Reference  0.5 (0.3-0.9) | Reference  0.6 (0.3-0.9) |
| Epidemic wave | 1  2  3 | 4.8±2.1 (2-8)  8.2±4.8 (2-21)  7.4±4.8 (2-21) | Reference  0.4 (0.2-0.6)  0.4 (0.2-0.7) |  |
| Variant | Wild type | 4.7±2.2 (2-8) | Reference | Reference |
|  | Beta  Alpha  Delta | 7.8±4.5 (2-21)  12.8±4.4 (8-17)  7.5±4.9 (2-21) | 0.4 (0.2-0.7)  0.2 (0.1-0.6)  0.4 (0.2-0.7) | 0.2 (0.1-0.6)  0.4 (0.2-0.6)  0.4 (0.2-0.7) |
| **Characteristics of the household member** | | | |  |
| Age group (years) | <5  5-12  13-18  19-39  40-59  ≥60 | 7.9±5.1 (2-21)  7.0±4.4 (2-21)  7.9±5.3 (2-21)  7.1±4.6 (3-19)  8.1±4.7 (2-20)  8.4±4.7 (2-17) | 0.8 (0.5-1.3)  Reference  0.8 (0.5-1.2)  1.0 (0.6-1.5)  0.8 (0.5-1.3)  0.8 (0.4-1.4) |  |
| Sex | Female  Male | 7.2±4.6 (2-21)  8.0±4.8 (2-21) | 1.2 (0.9-1.5)  Reference |  |
| HIV and viral load copies/ml | Uninfected  Infected <400  Infected ≥400  HIV and/or viral load unknown | 7.5±4.8 (2-21)  7.3±4.1 (3-17)  8.9±5.6 (3-17) | Reference  1.1 (0.7-1.9)  0.8 (0.4-1.6)  1.4 (0.7-2.9) |  |
| HIV and CD4+ T cell count/ml | Uninfected  Infected ≥200  Infected <200  HIV and/or CD4+ unknown | 7.5±4.8 (2-21)  7.5±4.5 (3-17)  14±0.0 (14-14)  6.3±4.1 (3-12) | Reference  0.8 (0.6-1.3)  0.4 (0.1-3.0)  1.4 (0.6-3.0) |  |

SD – Standard deviation ^a^Estimated using Weibull accelerated failure time regression adjusted for clustering by site and household. Samples were collected at 2 to 4 day intervals. Serial interval refers to the interval between first positive influenza result in the index case and the secondary case.

Additional factors evaluated but not found to be statistically significant include year, site, employment of index or contact, education level of index or contact, alcohol or smoking of index or contact, other underlying illness of index or contact, body mass index of index or contact, receipt of influenza vaccine of index or contact, number of people in household, number of rooms, crowding, smoking inside the house.

Supplementary table 5: Characteristics associated with being an index case (vs non index case) for household clusters of SARS-CoV-2 infection in a rural and an urban site, South Africa, 2020-2021^a^

|  |  | **Index case**  **n/N (%)** | **Univariate** | **Multivariable** |
| --- | --- | --- | --- | --- |
| **Variable** |  |  | **RR (95% CI)** | **aRR (95% CI)** |
| Site | Rural  Urban | 200/350 (57)  191/326 (59) | Reference  1.1 (0.8-1.4) |  |
| Age group (years) | <5  5-12  13-18  19-39  40-59  ≥60 | 27/67 (40)  96/189 (51)  79/127 (62)  103/149 (69)  56/89 (63)  30/55 (55) | Reference  1.5 (0.9-2.7)  2.4 (1.3-4.5)  3.3 (1.8-6.0)  2.5 (1.3-4.8)  1.8 (0.9-3.7) | Reference  1.5 (0.8-2.7)  2.4 (1.3-4.5)  3.1 (1.6-5.9)  2.2 (1.1-4.2)  1.6 (0.7-3.4) |
| Sex | Female  Male | 231/412 (56)  160/264 (61) | 0.8 (0.6-1.1)  Reference |  |
| HIV and viral load copies/ml | Uninfected  Infected <400  Infected ≥400  HIV and/or viral load unknown | 308/549 (56)  49/71 (69)  13/23 (57)  21/33 (64) | Reference  1.7 (1.0-3.0)  1.0 (0.4-2.4)  1.4 (0.7-2.9) | Reference  1.4 (0.8-2.5)  0.8 (0.3-2.0)  1.8 (0.9-3.6) |
| HIV and CD4+ T cell count/ml | Uninfected  Infected ≥200  Infected <200  HIV and/or CD4+ unknown | 308/549 (56)  54/85 (64)  6/7 (86)  23/35 (66) | Reference  1.4 (0.8-2.2)  4.7 (0.6-39.3)  1.5 (0.7-3.1) |  |
| Other underlying illness^b^ | No  Yes | 355/614 (58)  36/62 (58) | Reference  1.0 (0.6-1.7) |  |
| BMI^c^ | Underweight  Normal weight  Overweight  Obese | 26/53 (49)  195/350 (56)  76/125 (61)  93/145 (64) | 0.8 (0.4-1.4)  Reference  1.2 (0.8-1.9)  1.4 (1.0-2.1) | 1.6 (0.9-2.9)  Reference  1.5 (0.7-2.9)  2.0 (0.9-3.9) |

^a^Estimated using logistic regression adjusted for clustering by site and household ^b^Self-reported history of asthma, lung disease, heart disease, stroke, spinal cord injury, epilepsy, organ transplant, immunosuppressive therapy, organ transplantation, cancer, liver disease, renal disease or diabetes ^c^BMI=body mass index calculated using the formula (weight in kilograms)/(height in metres squared). We defined BMI categories as follows: underweight - age <18 years weight for age or BMI <-2 standard deviations of the World Health Organization (WHO) Child Growth Standards, age ≥18 years BMI <18.5kg/m2; overweight - age <18 years BMI >+1 and ≤+2 standard deviations of the WHO growth standards, age ≥18 years BMI ≥25 and <30kg/m2, obese – age <18 years BMI >+2 standard deviations of the WHO growth standards, age ≥18 years BMI ≥30 kg/m2

Supplementary figure 1: Location of rural (Agincourt) and urban (Jouberton) study sites in South Africa

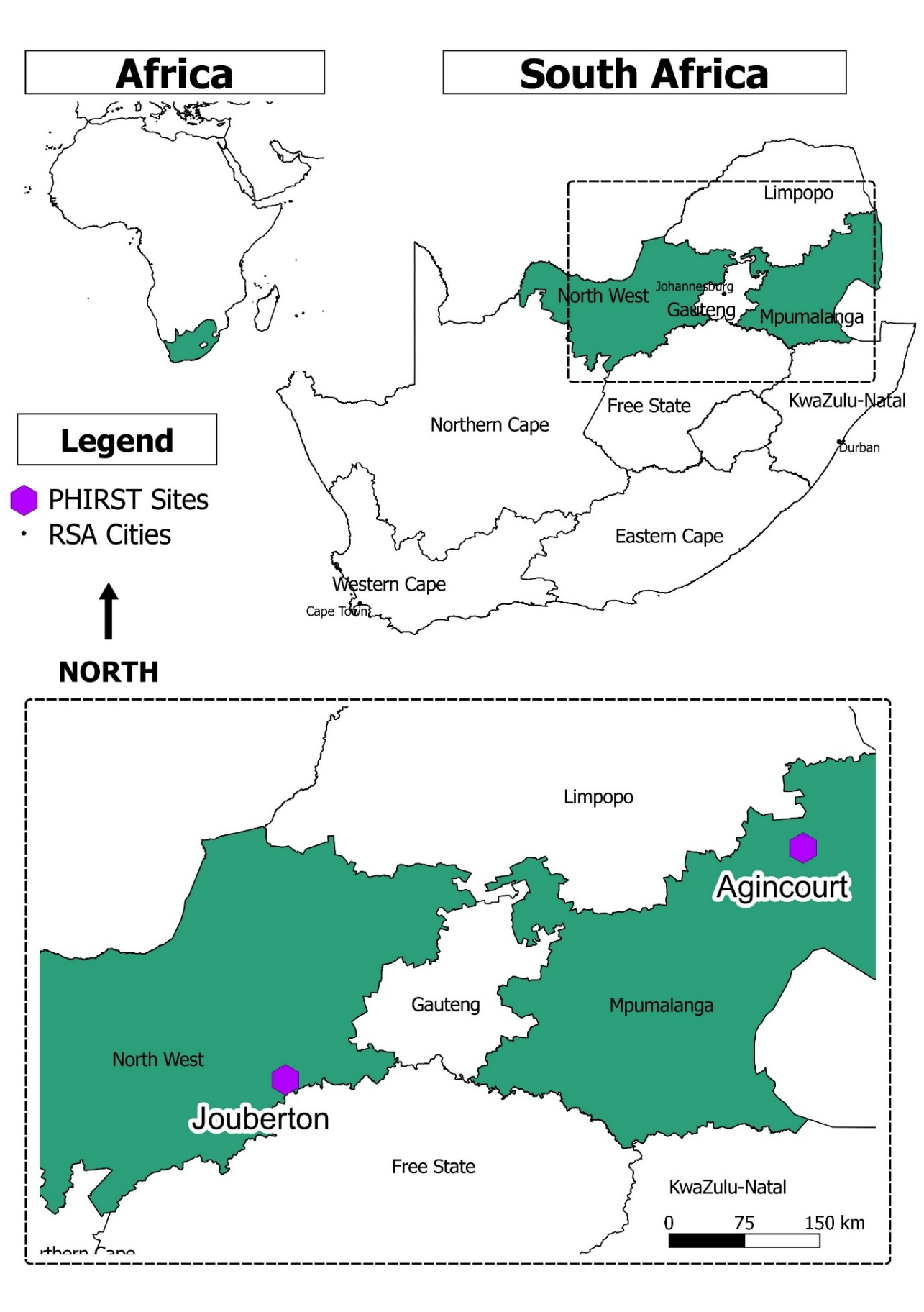

Supplementary figure 2: Methods for assigning episodes and clusters of infection.

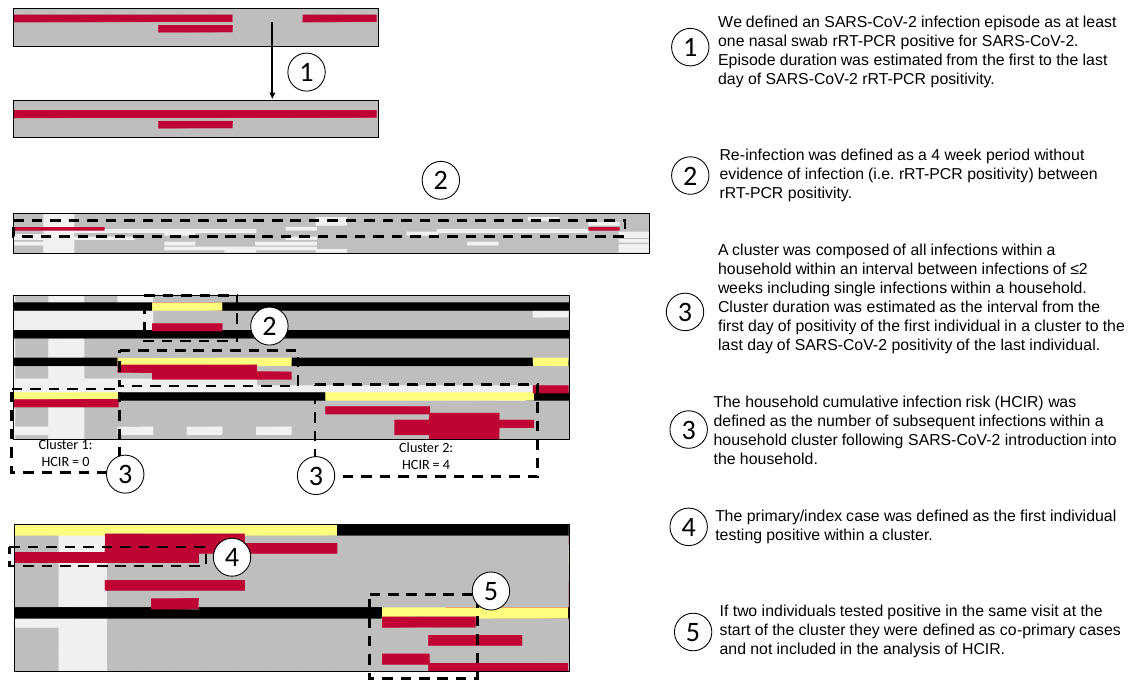

Figure legend

Red bar - Positive rRT-PCR

Dark Grey bar - Negative rRT-PCR

Light Grey bar - Missing swab

Black bar - No Household Cluster

Yellow bar - Household Cluster

Supplementary figure 3: Flow chart of individuals included in the study, a rural site and an urban site, South Africa, 2020-2021

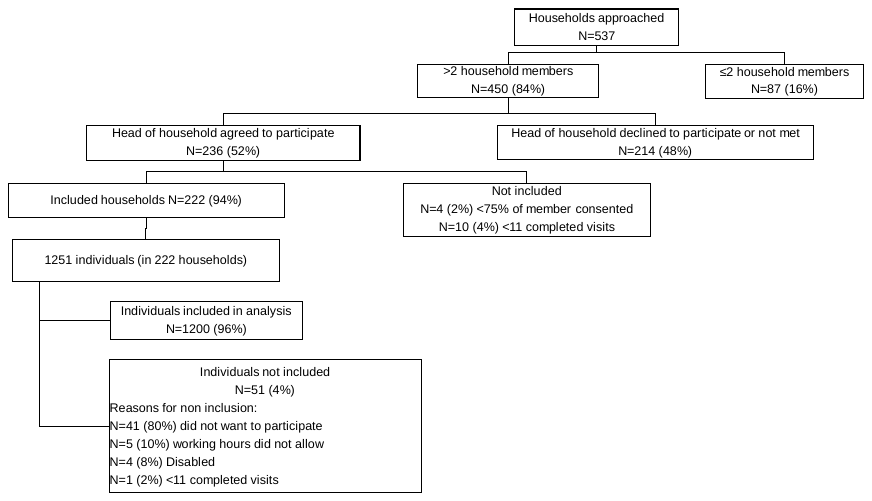

Supplementary figure 4: Proportion of individuals infected with SARS-CoV-2 at the end of follow up by method of diagnosis, age group and site at a rural and an urban site, South Africa, 2020-2021

a) Rural site	b) Urban site

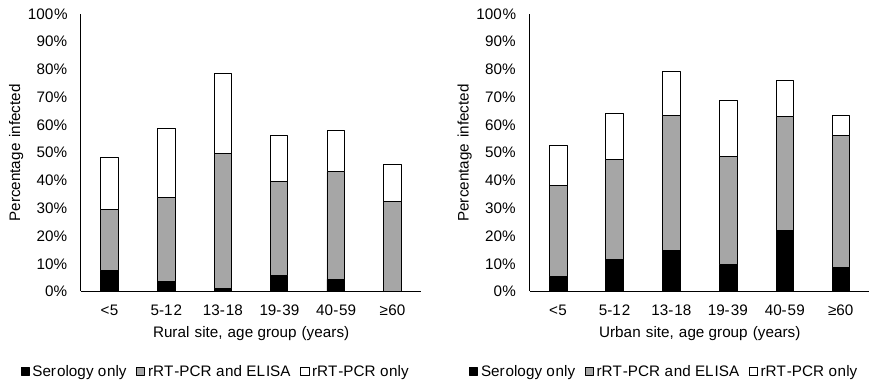

rRT-PCR- real time reverse transcription polymerase chain reaction

Supplementary figure 5: SARS-CoV-2 episodes of infection by SARS-CoV-2 variant at the rural site (a) and urban site (b) in 2020-2021.

Columns are individual follow up visits and rows are individual participants. The white, light grey and coloured horizontal lines each denote an individual with in a household. Each column indicates an individual follow up visit. Follow up visits are coloured white if no sample was tested, light grey if the sample tested negative for SARS-CoV-2 and coloured if the nasopharyngeal swab tested positive for SARS-CoV-2. Colours are green for non variant, red for Beta variant and blue for Alpha variant after imputation for missing data. Individuals in the same household make up sequential rows. A high resolution version of this figure has been provided separately.

A Rural site

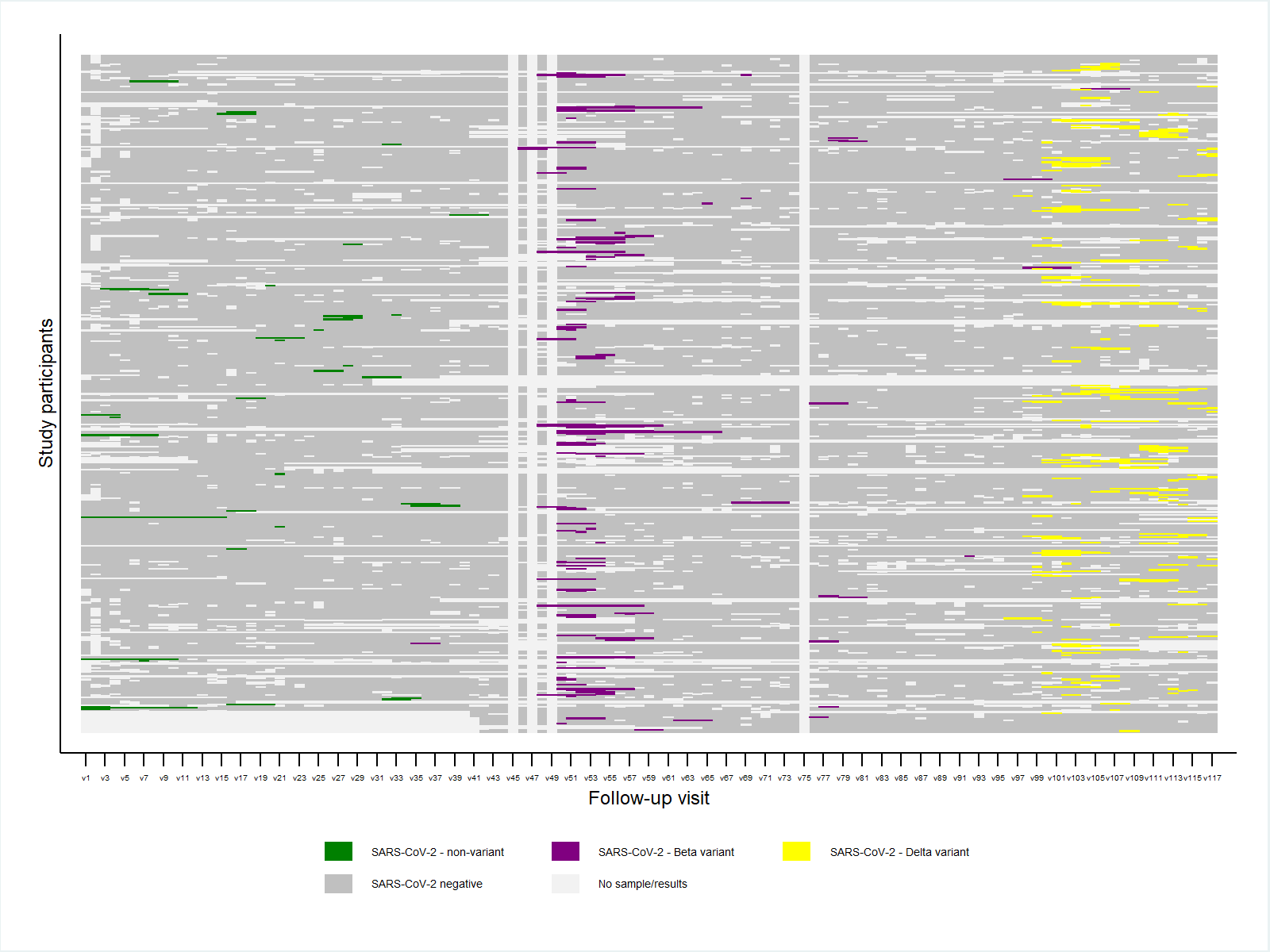

B) Urban site

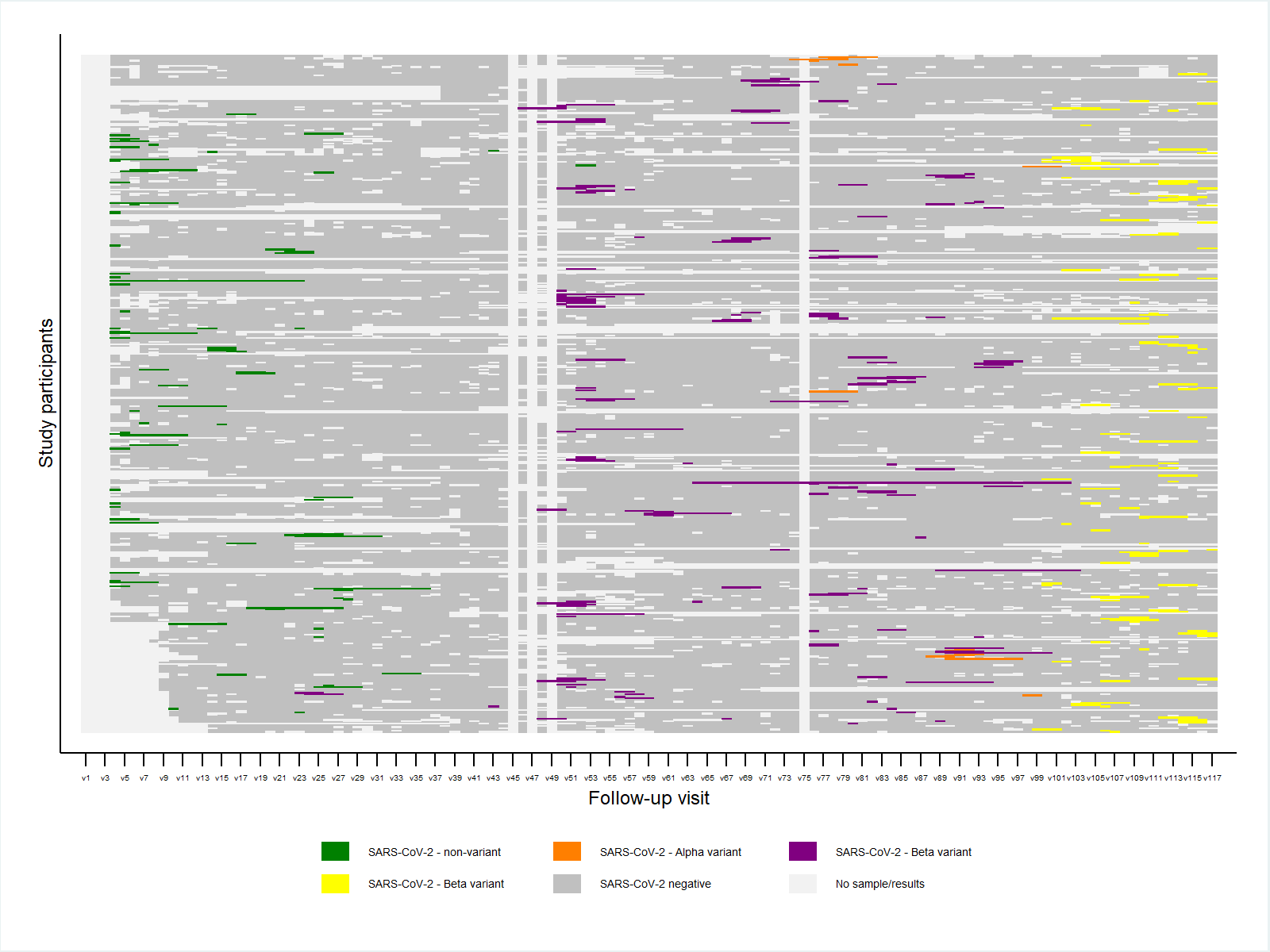

Supplementary figure 6: Epidemic curve of rRT-PCR-confirmed SARS-CoV-2 by non-variant or variant type at a rural and an urban site, South Africa, 2020-2021*

A) Rural site
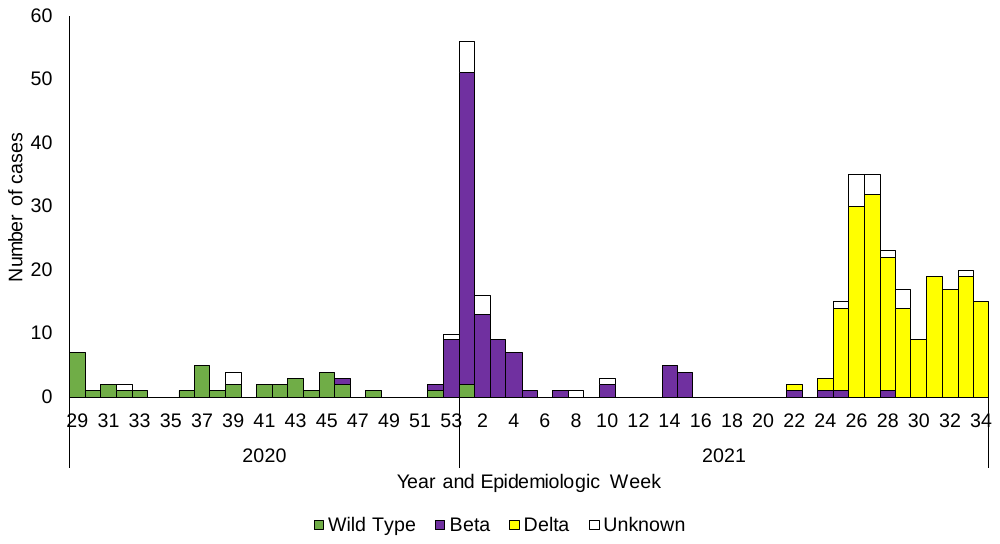

Wave 3

Wave 2

Wave 1

b) Urban site

Wave 3

Wave 2

Wave 1

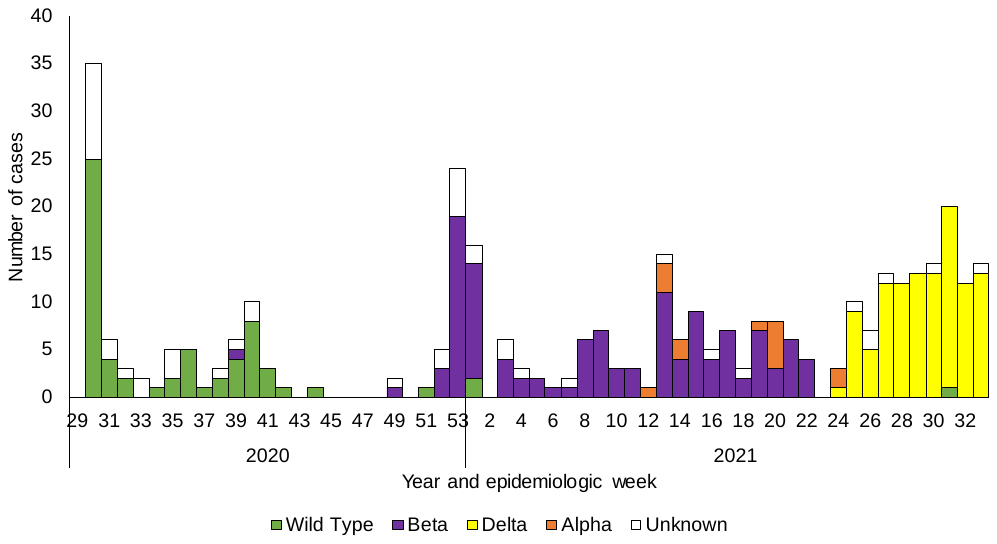

*Follow up began on 16 and 27 July 2020 at rural and urban site, respectively, vertical dashed lines indicates analysis cut off between first, second and third SARS-CoV-2 waves at each site

Supplementary figure 7: Interval between first SARS-CoV-2-positive real-time reverse transcription polymerase chain reaction (rRT-PCR) in the index case and first positive rRT-PCR in household contacts (generation interval), in a rural and an urban community, South Africa, 2020-2021 (n=212 case pairs, 198 included for generation interval estimation)

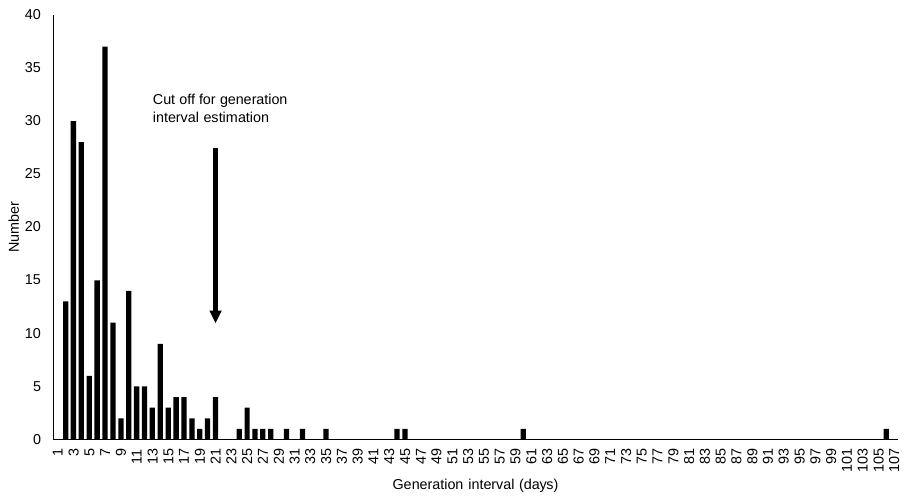
